# Parkinson’s disease genetic risk converges on Lewy body pathology-associated microglial states

**DOI:** 10.64898/2026.10.26.26364188

**Authors:** Raphael Kübler, Daniele Mattei, J. Oriol Narcis, Claudia De Sanctis, Kosei Hirata, Alice Buonfiglioli, Ariana Chriss, Mikaela Rosen Perez, Felix A. Crary, Emma L. Thorn, Alex Tokolyi, Tarek Khashan, Roy D. Missall, Beomjin Jang, BP Kailash, Nienke J. Mekkes, Melissa J. Nirenberg, Ruth H. Walker, David A. Knowles, Inge R. Holtman, Lot D. de Witte, John F. Crary, Joel Blanchard, Towfique Raj

**Author notes:** Correspondence: Towfique Raj.

## Abstract

Parkinson’s disease (PD) and related α-synucleinopathies are defined by the spread of Lewy body (LB) pathology across vulnerable brain regions, yet how substantia nigra (SN) myeloid cells respond remains unresolved. Using SOX10-based oligodendrocyte depletion and CD11b-based myeloid enrichment, we profiled 169,799 brain myeloid cells and nuclei from 96 donors spanning the LB continuum, including 86,427 from the SN, and integrated them with AMP PD for a total of 298,592 profiles. Across eight conserved states, α-synucleinopathy is associated with a bidirectional signature: upregulated glycolytic and proteostatic-stress programs alongside downregulated lysosomal and mitochondrial pathways. Microglia shift from P2RY12-high sensome cells toward an HSP90AA1-high proteostatic stress state, a transition that intensifies at late Braak LB stages. Immunohistochemistry confirmed that HSP60^+^ proteostatic microglia state proportions increase with SN neuron loss. A rare NUCKS1-high state (∼1% of myeloid cells), more abundant in late-stage disease, carried the strongest PD polygenic-risk loading. The PD-protective rs3732765-A allele, a microglial cis-eQTL that lowers *P2RY12* expression, was nominally associated with a higher proportion of P2RY12-high microglia and higher lysosomal program activity. These findings frame microglial involvement as structured and genetically tractable, nominating the *P2RY12* genetic axis and NUCKS1-high microglia for mechanistic studies.

## Introduction

Parkinson’s disease (PD) and dementia with Lewy bodies (DLB) are defined by the spread of α-synuclein pathology through vulnerable brain regions, culminating in the degeneration of substantia nigra (SN) dopaminergic neurons and the motor features of PD^1,2^. Genetic evidence, however, increasingly implicates brain myeloid cells: PD risk variants are enriched in microglial regulatory elements, and myeloid-expressed genes falling into the mitochondrial and proteolysosomal pathways are dysregulated by disease^3,4^. Microglia rapidly remodel their transcriptional programs in response to protein aggregates and neuronal injury^5–7^, yet whether this capacity is preserved, exhausted, or maladaptive in the degenerating human SN remains unknown.

Resolving this question requires deep transcriptomic sampling of SN myeloid cells, and here prior studies have been limited. Across five recent studies, fewer than 70,000 SN myeloid nuclei have been recovered in total^8–12^, with individual studies capturing 4,000 to 30,000, insufficient to resolve state heterogeneity, quantify abundance shifts across disease stages, or power genetic analyses. Deep myeloid profiling has been achieved in Alzheimer’s disease through enrichment-based approaches (PsychAD)^13^, but PD studies have relied on standard nuclear preparations that yield only 5-15% microglia. The Accelerating Medicines Partnership Parkinson’s Disease (AMP PD) recently described microglial remodeling along Braak Lewy body (LB) stages using brainstem, pallidal, and cortical nuclei^6^, but their resource did not include the SN. Hence, no existing PD resource combines myeloid enrichment, cross-cohort harmonization, and the donor scale needed for systematic analysis. As a result, the field lacks a unified molecular taxonomy of human SN myeloid cells and a clear view of the transcriptional programs engaged by α-synuclein pathology.

We addressed this gap with the myeloid arm of an Atlas of Integrative Omics of Neuroimmunity in α-synucleinopathy (AION-myeloid), an integrated atlas of 169,799 human brain myeloid profiles from 96 unique donors spanning the full spectrum of LB pathology. Using SOX10-based oligodendrocyte depletion and CD11b-based myeloid enrichment, we recovered 86,427 substantia nigra myeloid transcriptional profiles from α-synucleinopathy and neurotypical control donors, exceeding all prior PD SN datasets combined. Additional sampling from amygdala and cortex (83,372 transcriptional profiles), together with integration of 128,793 myeloid nuclei from the AMP PD consortium^6^, provides coverage of brain regions variably affected across Braak LB stages. Together, these data support a systematic molecular dissection of SN myeloid biology in PD: defining a consensus cell state taxonomy, tracking state-specific abundance shifts across the LB pathology continuum, identifying transcriptional programs that scale with pathological progression, and linking myeloid states and gene programs to PD genetic risk.

## Results

### Taxonomy of α-synucleinopathy brain myeloid cells

AION-myeloid profiles human brain myeloid cells with two complementary enrichment strategies: SOX10-depleted nuclei isolated by fluorescence-activated nuclei sorting (FANS) from cortex (CTX), amygdala (AMY) and substantia nigra (SN), and CD11b-enriched cells isolated by magnetic-activated cell sorting (MACS) from CTX and SN. For the abundance and transcriptional analyses, we restricted the cohort to the α-synucleinopathy spectrum. To avoid confounding by unrelated pathologies, we removed donors with progressive supranuclear palsy (PSP), amyotrophic lateral sclerosis (ALS) or Alzheimer’s disease neuropathologic change (ADNC), retaining 36,214 of 37,310 SOX10-depleted myeloid nuclei and 133,585 CD11b-enriched myeloid cells. AION-myeloid was designed to extend existing PD single-cell resources such as AMP PD by adding myeloid enrichment, regions not covered in the AMP PD data used here (AMY, SN) and matched donor genotypes. We therefore integrated it with 128,793 AMP PD myeloid nuclei from CTX, globus pallidus interna (GPI) and dorsal motor nucleus of the vagus (DMNX). This yielded a harmonized dataset of 298,592 myeloid cells and nuclei from 96 AION-myeloid and 97 AMP PD donors (**Table 1**, **Fig. 1a,c**), with brain region included as a covariate to account for uneven regional coverage across sources. For the genetic analyses, we retained PSP, ALS and ADNC donors to preserve power, using all 113 genotyped AION-myeloid donors (72.6% European genetic ancestry; 260,844 myeloid nuclei and cells) across brain regions; AMP PD was excluded because donor genotypes were not available.

**Figure 1.**
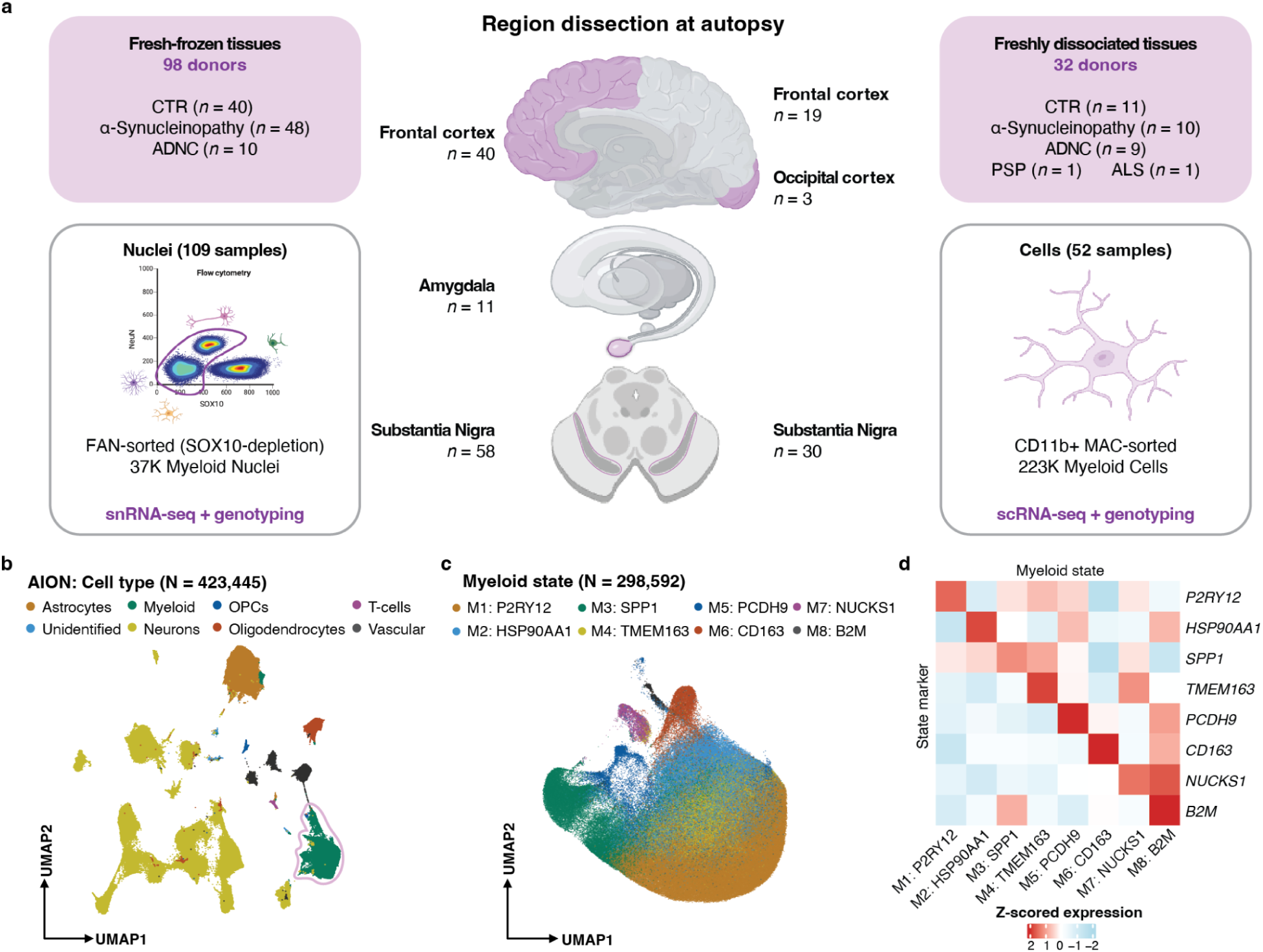
**(a)** Overview of the study design. Human postmortem cortex (CTX), amygdala (AMY), and substantia nigra (SN) were profiled by SOX10-depleted single-nucleus RNA-seq and CD11b-enriched single-cell RNA-seq across 113 unique donors. **(b)** UMAP of in-house nuclei colored by cell-type annotations. The myeloid population is marked with a light-pink outline. **(c)** Joint CONCORD UMAP embedding (**Methods**) of all myeloid cells and nuclei (AION-myeloid + AMP PD), showing harmonized clustering across datasets. **(d)** Heatmap of top marker pseudobulked gene expression defining the eight myeloid states: P2RY12-high, SPP1-high, HSP90AA1-high, TMEM163-high, PCDH9-high, CD163-high, NUCKS1-high, and B2M-high. GPI = Globus Pallidus Internus, DMNX = Dorsal Motor Nucleus of the Vagus. Donor and sample numbers in **(a)** are pre-QC; post-QC numbers are given in **Table 1**.

**Table 1.** Cohort overview. Demographic, neuropathological and single-cell profiling summary of the 113 post-mortem donors in the myeloid cell/nucleus atlas, grouped by primary neuropathological diagnosis. Continuous variables are given as median [IQR] and categorical variables as n (%). The *ADNC* column denotes donors whose primary diagnosis is Alzheimer’s disease-type pathology, whereas the *ADNC (NIA-AA)* rows report the NIA-AA co-pathology score across all donors. *APOE* ε4 carriers are donors with ≥1 ε4 allele; recovered cells/nuclei are post-QC myeloid counts. Braak LB, Braak Lewy body stage (binned); PMI, post-mortem interval; IQR, interquartile range; snRNA-seq, single-nucleus RNA sequencing; scRNA-seq, single-cell RNA sequencing; MG, microglia; PSP, progressive supranuclear palsy; ALS, amyotrophic lateral sclerosis; contributing brain banks: NPBB, Neuropathology Brain Bank at Mount Sinai; NBB, Netherlands Brain Bank.

|  | Control | Synucleinopathy | ADNC | Other<br>(PSP/ALS) | Total |
| --- | --- | --- | --- | --- | --- |
| n donors | 45 | 51 | 15 | 2 | 113 |
| Female, n (%) | 21 (47) | 12 (24) | 6 (40) | 1 (50) | 40 (35) |
| Age at death, median [IQR] | 78 [65-87] | 78 [74-86] | 74 [73-80] | 72 [71-74] | 77 [73-85] |
| PMI (h), median [IQR] | 11 [6-20] | 12 [6-22] | 18 [12-22] | 17 [17-18] | 12 [6-22] |
| Cohort, NPBB / NBB | 25 / 20 | 31 / 20 | 15 / 0 | 2 / 0 | 73 / 40 |
| **Braak LB** |  |  |  |  |  |
| Control | 45 | 0 | 15 | 2 | 62 |
| Intermediate | 0 | 11 | 0 | 0 | 11 |
| Late | 0 | 40 | 0 | 0 | 40 |
| **ADNC (NIA-AA)** |  |  |  |  |  |
| Not AD | 20 | 14 | 3 | 1 | 38 |
| Low | 15 | 12 | 4 | 0 | 31 |
| Intermediate | 3 | 13 | 4 | 0 | 20 |
| High | 0 | 3 | 4 | 0 | 7 |
| Not assessed | 7 | 9 | 0 | 0 | 16 |
| APOE $\epsilon 4$ carriers, n (%) | 11 (24) | 21 (41) | 2 (13) | 1 (50) | 35 (31) |
| Profiled - snRNA-seq (donors) | 39 | 48 | 10 | 0 | 97 |
| Profiled - scRNA-seq (donors) | 11 | 10 | 9 | 2 | 32 |
| Both modalities (donors) | 5 | 7 | 4 | 0 | 16 |
| **Cells/nuclei recovered, n** |  |  |  |  |  |
| AION-myeloid (nuclei) | 13,820 | 22,394 | 1,096 | 0 | 37,310 |
| AION-myeloid (cells) | 82,394 | 51,191 | 54,272 | 35,677 | 223,534 |

Unsupervised clustering and marker-based annotation revealed seven major cell-type clusters in AION (nuclei; **Fig. 1b**; **Supplementary Fig. 1a**): neurons, astrocytes, myeloid cells, oligodendrocyte precursor cells (OPCs), oligodendrocytes, T-cells, and vascular cells. We then performed in *silico* subsetting and unsupervised clustering of the myeloid populations in AION (nuclei). This yielded AION-myeloid (nuclei), a dataset comprising eight myeloid states, each defined by distinct transcriptional programs and named for a representative marker gene (**Fig. 1d**, **Supplementary Fig. 1b**). These included seven states of microglial identity (P2RY12-high, SPP1-high, HSP90AA1-high, TMEM163-high, PCDH9-high, NUCKS1-high, and B2M-high), and one state of border-associated macrophages (CD163-high). Several of the top state markers are established PD risk genes, each expressed in a largely state-restricted pattern (**Supplementary Fig. 1c**), a partitioning we revisit when assessing polygenic risk across states.

These states were reproducible across cohorts and their identity was robust to modality. Using the contrastive learning method CONCORD^14^ to propagate state labels across datasets, Pearson correlation of each state’s top 50 markers indicated that the states replicated across our myeloid cohorts (**Supplementary Fig. 2i-k**). In a matched-donor subset of AION-myeloid, a linear mixed model (see **Methods**) showed that nuclei and cells differed only in genes from cytoplasmic pathways, the compartment absent from nuclei (**Supplementary Fig. 3**).

Over-representation analysis (ORA) of Gene Ontology Biological Process (GO:BP) terms across the top 50 marker genes per state assigned seven of the eight states to previously described myeloid transcriptional programs: P2RY12-high (M1) corresponds to neuronal surveillance (i.e., sensome functions)^15^; HSP90AA1-high (M2) to proteostatic stress and the adaptive response to unfolded proteins^16^; SPP1-high (M3) to MHC-II and ribosomal enrichment associated with phagocytic functions^17^; TMEM163-high (M4) to cell migratory and inflammatory programs^18^; PCDH9-high (M5) to synapse regulation^19^; CD163-high (M6) to perivascular macrophages and immune/inflammatory signaling^20,21^; and B2M-high (M8) to phagocytosis-induced neuroinflammation^22^ (**Supplementary Fig. 4**). NUCKS1-high (M7) microglia are the exception: they co-express markers spanning sensome (*P2RY12*), phagocytic (*SPP1*), and inflammatory (*TMEM163*) states (**Fig. 1d)** and show no enrichment for any canonical biological pathway (**Supplementary Fig. 4**), consistent with a hybrid or transitional identity rather than a discrete transcriptional program. Across our three cohorts and both modalities (nuclei and cells), the NUCKS1-high state accounts for 0.5–1.3% of myeloid cells (**Supplementary Fig. 2e**). Although small, it remains a discrete, reproducible population rather than a technical artifact: it clusters stably (**Supplementary Fig. 2i-k**), is de-enriched for apoptosis and cell-death markers (**Supplementary Fig. 5**), and its abundance largely is unrelated to technical covariates (**Supplementary Fig. 6**). To our knowledge, no existing literature describes the function of NUCKS1-high microglia in health or disease.

To look for a program below genome-wide resolution, we used the state’s defining marker as a lead. NUCKS1 is a chromatin-associated factor involved in homologous recombination, DNA repair and cell-cycle progression^23–25^, with no described role in microglia. Targeted ORA of the 50 top state markers against curated cell-cycle gene sets showed that NUCKS1-high microglia were the only state enriched for a synthesis (S)-phase signature (**Supplementary Fig. 7**), pointing to a proliferative or DNA-replicative component. We next tested whether the abundance of this population shifts with disease stage.

### Sensome microglia are depleted while proteostatic stress and S-phase microglia expand in α-synucleinopathy

We first assessed whether α-synucleinopathy alters myeloid state composition across brain regions. We modeled cell-state abundance with crumblr^26^, which applies a variance-stabilizing transformation to cell-type proportions with precision weights that account for the compositional structure of the data. We fit linear mixed models with dream^27^, adjusting for age, sex, brain region, post-mortem interval and sample-level quality metrics, with a random intercept for donor (**Methods**). Braak LB stages were binned into control (Braak LB stage 0), intermediate (Braak LB stages 1-4), and late (Braak LB stages 5-6).

We then tested intermediate-vs-control and late-vs-control contrasts separately. Both contrasts recovered largely the same directional shifts across all three datasets, with effect sizes generally larger at the late stage: P2RY12-high microglia were depleted, while HSP90AA1-high and NUCKS1-high microglia expanded. These shifts reached significance in the merged late-vs-control analysis (P2RY12-high: logFC = −0.40, FDR = 0.092; HSP90AA1-high: logFC = 0.58, FDR = 0.031; NUCKS1-high: logFC = 0.56, FDR = 0.011; **Fig. 2a-c**; **Supplementary Fig. 8**). In the merged dataset, the intermediate-vs-control contrast was directionally concordant, with smaller effects that did not reach significance (all FDR ≥ 0.1). This is consistent with a milder shift at the intermediate stage and fewer intermediate donors, although HSP90AA1-high expansion was already detectable at the intermediate stage in AMP PD. Other states showed variable or smaller effects, including an expansion of SPP1-high microglia in AMP PD and modest depletion of PCDH9-high and B2M-high microglia in the merged late-vs-control analysis (**Fig. 2a-c**; **Supplementary Fig. 8**).

**Figure 2.**
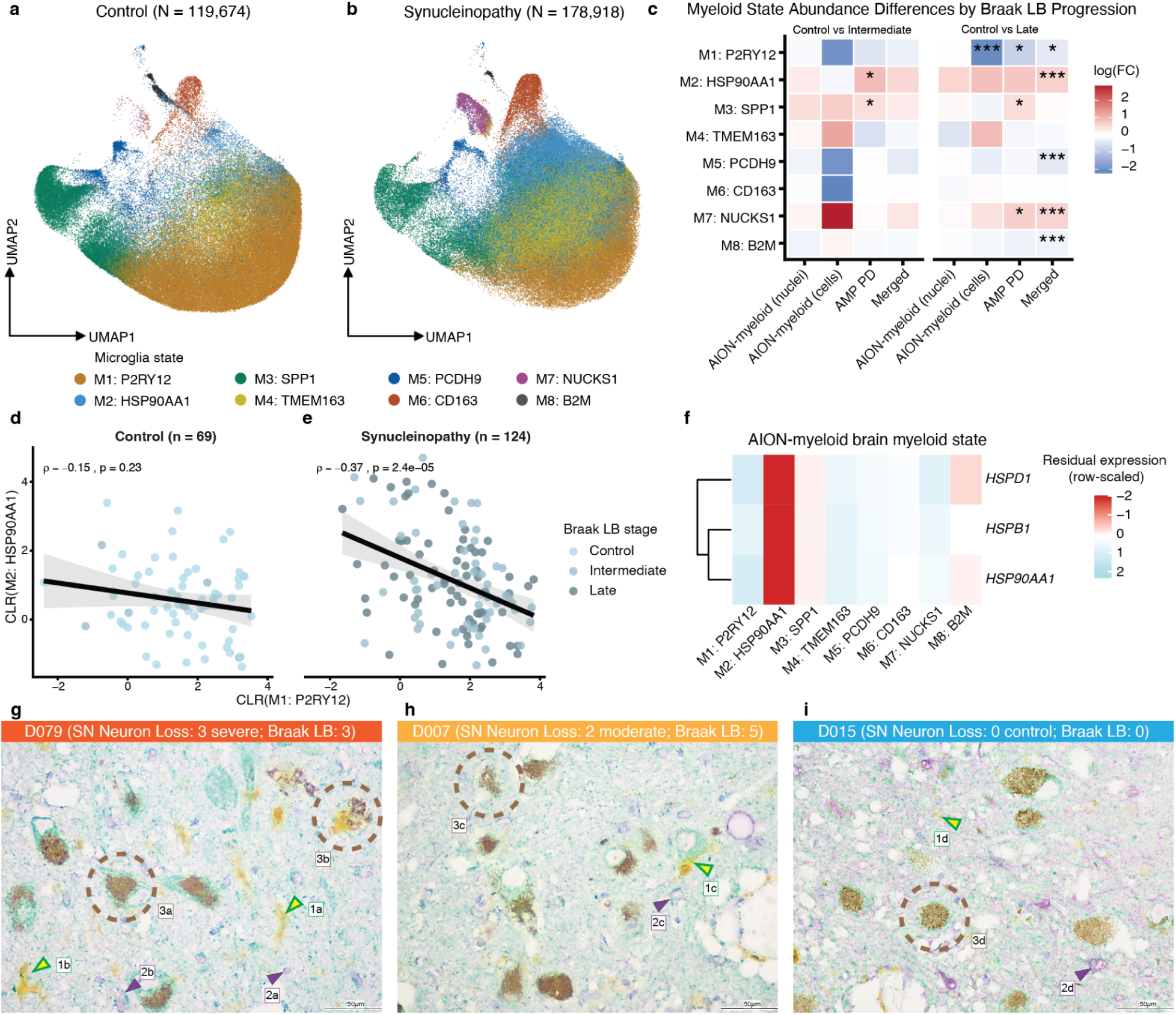
**(a-b)** Joint CONCORD UMAP embeddings (**Methods**) of control and α-synucleinopathy donors showing redistribution of P2RY12-high and HSP90AA1-high microglia. **(c)** Cohort-integrated abundance modeling showing consistent depletion of P2RY12-high microglia and enrichment of HSP90AA1-high and NUCKS1-high microglia across Braak LB stages (Control = Braak LB stage of 0; Intermediate = Braak LB stage of 1-4; Late = Braak LB stage of 5-6). ***FDR < 0.01, **FDR < 0.05, *FDR < 0.1. **(d-e)** Sample-level negative correlation between P2RY12-high and HSP90AA1-high microglial abundances, stratified by pathology group. **(f)** Heatmap of Heat Shock Protein (HSP) gene expression in our AION-myeloid data per state. *HSPD1* encodes for HSP60. **(g-i)** Representative triple chromogenic IHC stainings from three donors (total IHC n = 12). HSP60 in green, IBA1 in yellow, and ALDH1L1 in purple. 1a-d = IBA1^+^/HSP60^+^ cell, 2a-d = ALDH1L1^+^/HSP60^+^ cell, 3a,d = intact Neuromelanin^+^ signal. 3b,c = dispersed Neuromelanin^+^ signal.

If α-synucleinopathy drives sensome microglia into the proteostatic-stress state, the two populations should be inversely coupled across donors, and more strongly so in disease. Consistent with this, P2RY12-high and HSP90AA1-high proportions were negatively correlated at the donor level, with the strength of the association between the two states differing by disease status: no significant correlation in controls (Spearman ρ = −0.15, *P* = 0.23) and moderate in α-synucleinopathy donors (ρ = −0.37, *P* = 2.4 × 10⁻⁵; **Fig. 2d-e**). This suggests microglial compositional remodeling occurs predominantly within the disease state.

To validate this shift at the protein level, we performed triple chromogenic immunohistochemistry (IHC; IBA1/HSP60/ALDH1L1) on SN from twelve Neuropathology Brain Bank & Research CoRE at Mount Sinai (NPBB) donors (**Fig. 2g-i**; **Supplementary Fig. 9**). HSP60 was chosen as the readout because its gene, *HSPD1*, is among the most strongly upregulated genes in the HSP90AA1-high state (**Fig. 2f**), and because HSP60 is a mitochondrial chaperonin that, when released, acts as a danger signal to the innate immune system^28^. HSP60 co-localized with IBA1+ microglia, and the proportion of IBA1+ microglia co-expressing HSP60 increased with SN neuron-loss severity (**Supplementary Fig. 9**). In the eleven donors with both IHC and sn/scRNA-seq data from SN (one slide excluded because of tissue damage), the proportion of proteostatic stress microglia rose with neuron loss in both IHC (IBA1^+^/HSP60^+^; 17% with no SN neuron loss to 41% with severe SN neuron loss, *P* = 0.019) and sn/scRNA-seq (HSP90AA1-high microglia; 11% to 45%, *P* = 0.001), with no detectable difference between the two methods (*P* = 0.39; **Supplementary Fig. 9**). Together, these results provide an orthogonal, protein-level confirmation that the proteostatic-stress (HSP90AA1-high) state expands with dopaminergic neuron loss in the degenerating SN.

After adjusting for LB pathology, myeloid state abundance differed markedly by brain region, whereas sex and age effects were modest (**Supplementary Figs. 10-11**). Regionally, HSP90AA1-high microglia were enriched in the DMNX (logFC = 0.59, *P* = 4.4 × 10⁻⁹) and GPI (logFC = 0.30, *P* = 0.004) and depleted in cortex (logFC = −0.68, *P* = 3.7 × 10⁻¹⁵; AMP PD, each region vs. all others), with concordant estimates in the merged dataset (**Supplementary Fig. 10a-b**). By sex, males (n = 73) showed higher HSP90AA1-high and females higher TMEM163-high abundance. The direction was consistent across all three datasets and the merged analysis, but the effects reached only nominal significance (HSP90AA1-high,

AION-myeloid (nuclei): logFC = 0.73, *P* = 0.048; TMEM163-high, merged: logFC = −0.42, *P* = 0.018) and did not survive FDR correction (**Supplementary Fig. 11a-b**). After adjusting for pathology, age was not associated with HSP90AA1-high or P2RY12-high abundance in any dataset (all P ≥ 0.1; **Supplementary Fig. 11c-d**). This suggests that apparent age-related shifts in these states largely reflect the co-accumulation of Lewy pathology with age. The strongest age association was a decrease in B2M-high microglia (merged: logFC = −0.12, *P* = 0.004). No sex or age association survived FDR correction.

Together, these data show a reorganization of the myeloid compartment in α-synucleinopathy: the balance shifts away from sensome (P2RY12-high) microglia that survey the parenchyma toward HSP90AA1-high microglia engaged in the heat-shock response, while the rare NUCKS1-high population expands. This shift is stronger in late Braak LB stage donors, is reproduced across datasets, and sits on a regional baseline in which HSP90AA1-high microglia are most abundant in the early-affected DMNX and least abundant in cortex.

### Microglia in α-synucleinopathy exhibit coordinated metabolic-proteostatic remodeling and lysosomal decline

To characterize transcriptional changes associated with α-synuclein pathology, we performed pseudobulk differential gene expression (DGE) analysis using a linear mixed model^27^ with donor as a random effect, comparing α-synucleinopathy to control samples at both myeloid-wide and state-specific levels, adjusting for age, sex, post-mortem interval, brain region, dataset, and additional technical covariates (**Methods**). Myeloid-wide analysis identified 105 upregulated and 89 downregulated genes (FDR < 0.05, |log_2_FC| > 0.5; **Fig. 3a**, **Supplementary Table 1**). ORA of the top differentially expressed genes at FDR < 0.1 revealed a bidirectional pattern (**Fig. 3b**). Upregulated genes were enriched for glycolytic metabolism and Heat Shock Response (HSR); downregulated genes were enriched for lysosomal modules, electron transport chain (ETC) pathways, and interferon signaling. Targeted module score (i.e., the z-scored mean of a pathway’s genes) testing confirmed these shifts at the donor level: HSR and F6P Phosphorylation pathway genes increased in expression across pathology and Braak LB stages (*P* < 0.01; **Fig. 3c,d**, **Supplementary Fig. 12a-b**).

**Figure 3.**
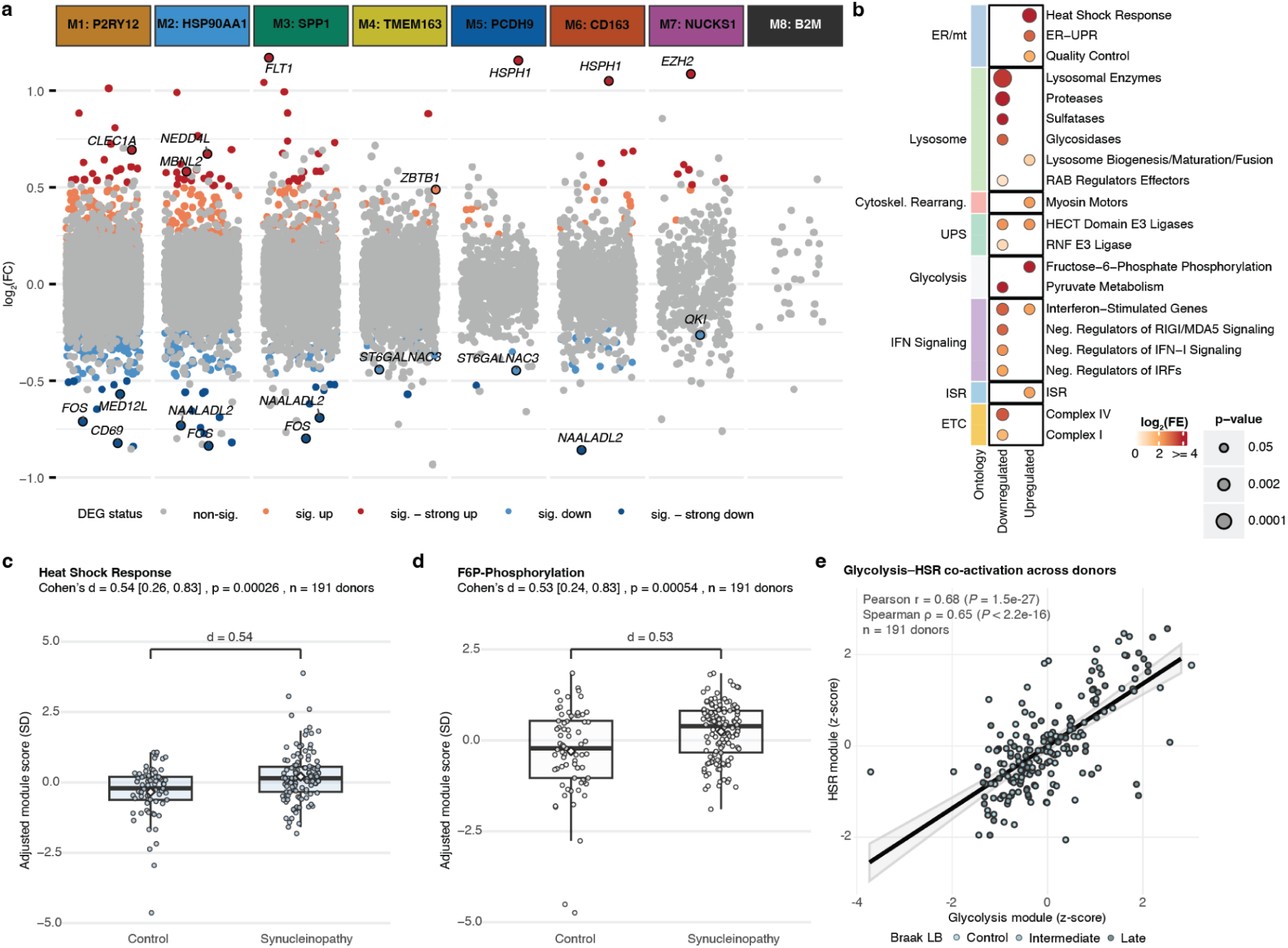
**(a)** Volcano plot of differentially expressed genes (DEGs) between α-synucleinopathy and control donors per myeloid state (FDR < 0.05, |log_2_FC| > 0.5). The two top differentially expressed genes (one down, one up) and PD risk gene hits are highlighted. **(b)** Pathway enrichment (ORA) of global DEGs (FDR < 0.1) across regions showing upregulation of glycolysis and HSR programs and downregulation of lysosomal, ETC, and IFN signaling programs. Dot sizes indicate nominal *P*-value of enrichment. **(c-d)** Donor-level module score (i.e., z-scored mean of that pathway’s genes) of Heat Shock Response (HSR) programs and F6P-Phosphorylation in α-synucleinopathy vs control donors. **(e)** Scatter plot showing sample-level co-activation of glycolytic and HSR programs (Pearson *r* = 0.68; n = 191 of 193 donors; 2 donors lacked PMI and were excluded from the covariate-adjusted models). Two donors were excluded in the analyses due to missing PMI (final n = 191).

We previously^29^ positioned metabolic reprogramming downstream of proteostatic stress in peripheral monocytes from PD patients. We applied the same internally curated gene sets to brain myeloid states, targeting PD-relevant cellular functions and resolving discrete functional programs more finely than broad GO biological-process terms. We first tested whether HSR and glycolytic pathway expression were correlated across donors. Indeed, their activity was tightly linked (Pearson *r* = 0.68, *P* = 1.5 × 10^−2^^7^; **Fig. 3e**).

To assess whether state-specific contributions drove these global myeloid DGE patterns, we fit a linear mixed model on state-subsampled cells and nuclei and performed ORA on the state-level DEGs against the curated functional panel (**Supplementary Fig. 13a-b**, **Supplementary Table 2**). Because this panel is a small, hypothesis-driven set rather than a discovery database, we report nominal p-values.

Enrichment was concentrated in the three states carrying the largest DE signal (P2RY12-high, HSP90AA1-high, SPP1-high microglia). SPP1-high microglia showed the strongest upregulation, a coordinated stress and innate-immune axis spanning the mitochondrial/ER heat shock response (HSR; *DNAJA1*, *HSPD1*; *P* = 0.004) and cytosolic nucleic-acid sensing (dsRNA/DNA sensors and effectors; *TRAF3*, *STAU2*; *P* = 0.013), while sensome P2RY12-high microglia upregulated interferon-stimulated genes (*ZC3HAV1*, *NLRC5*; *P* = 0.012). Upregulation of HECT-domain E3 ligases (ubiquitin-proteasome system) was a recurring feature across the P2RY12-, HSP90AA1-, SPP1-, and CD163-high states, though below nominal significance. Downregulation converged on clearance and cytoskeletal machinery: CD163-high perivascular macrophages downregulated lysosome biogenesis/maturation/fusion (*STX8*, *SUMF1*; *P* = 0.014) alongside weaker reductions in myosin motors and RAB effectors, and P2RY12-high microglia downregulated actin-binding proteins (*CFL1*, *AIF1*; *P* = 0.026); SPP1- and HSP90AA1-high microglia showed weaker, trafficking- and protease-oriented lysosomal downregulation (RAB effectors and lysosomal proteases/enzymes, respectively). The remaining low-DE states (TMEM163-high, PCDH9-high) produced only isolated enrichments, and NUCKS1-high microglia yielded too few DEGs to overlap the panel, showing no enrichment in either direction.

We next asked whether microglial remodeling in α-synucleinopathy reflects a brain-specific or a systemic myeloid response. To this end, we performed ORA using published DEG signatures from peripheral monocytes of idiopathic PD patients (Mattei *et al.*^29^). Pathway directionality was conserved: glycolysis and HSR programs were upregulated, while lysosomal pathways were downregulated, in both brain myeloid cells and peripheral monocytes (Pearson’s r = 0.49 for upregulated pathways; Pearson’s r = 0.35 for downregulated pathways; **Supplementary Fig. 14**). This convergence suggests a shared myeloid stress signature operating across tissue compartments in PD.

Together, these findings indicate that α-synucleinopathy is associated with a coordinated metabolic and proteostatic shift in brain myeloid cells. The concurrent downregulation of ETC genes alongside upregulation of glycolytic and HSR programs, whose activities were tightly correlated across donors, is consistent with a metabolic adaptation to compromised mitochondrial homeostasis and proteostatic stress, accompanied by a shift toward glycolysis. This pattern parallels the coupled proteostatic and metabolic reprogramming reported in peripheral PD monocytes, where reduced mitochondrial function is evident both transcriptionally and functionally, and its directionality was conserved across compartments here. Suppression of lysosomal and trafficking pathways, concentrated in phagocytic populations, further points to a reduced capacity for aggregate clearance. The state-resolved analysis, though limited by sparse per-state signal, was broadly consistent with this global pattern rather than pointing to a single dominant state: the proteostatic and innate-sensing arm was strongest in SPP1-high microglia, lysosomal and trafficking suppression in CD163-high perivascular macrophages, and an interferon-stimulated signature in sensome P2RY12-high microglia.

### Heat-shock and lysosomal gene programs correlate with Lewy body pathology

Previously, Marshe *et al*.^30^ noted that cluster-based analyses can impose sharp boundaries on microglial variation that is partly continuous. To complement our state-level analysis, we used Spectra^31^ to model microglial expression as a set of additive gene programs rather than discrete clusters. Spectra is a pathway-guided factorization method that identifies co-regulated transcriptional modules, here referred to as Spectra factors. Factors were labeled by their seeding gene sets; ORA on top-loading genes was used to annotate factors and to characterize *de novo* factors not anchored to prior knowledge. Factor activity was aggregated per donor and compared between α-synucleinopathy and control donors using mixed models adjusting for age, sex, PMI and data type (i.e., nuclei and cells; **Methods**). Of 20 factors tested, 7 differed between groups (FDR < 0.1). Six were lower in α-synucleinopathy: lysosomal enzymes, nucleases and proteases, mitochondrial quality control, Complex V (ATP synthase) and, notably given prevailing activation models, pathogen recognition (adjusted differences −0.11 to −0.21 SD; FDR < 0.05; **Fig. 4a-f**). The HSR factor was the only factor higher in α-synucleinopathy (+0.32 SD; FDR = 0.076; **Fig. 4g**; **Supplementary Table 4**).

**Figure 4.**
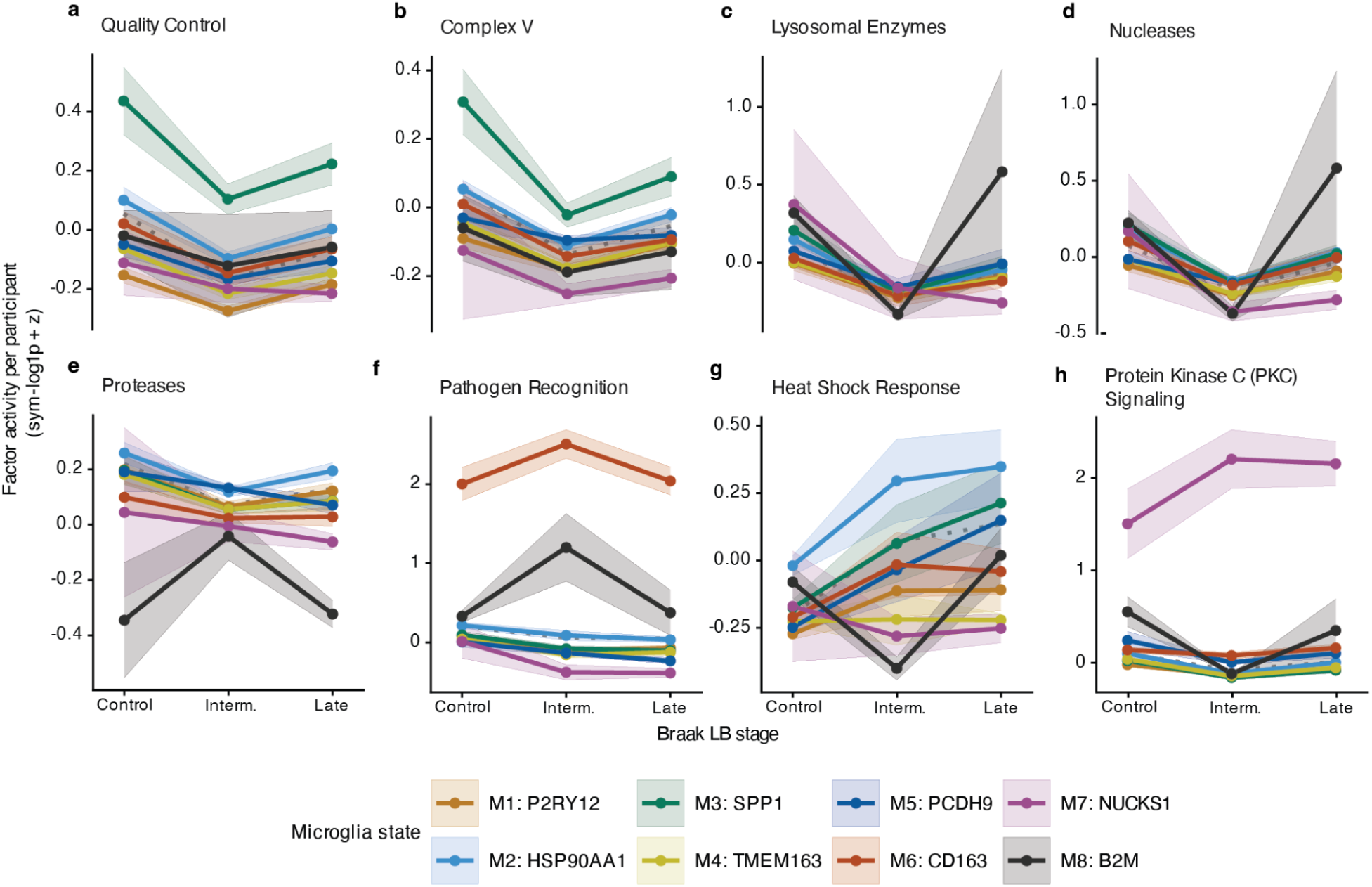
Spectra factor scores (donor-level collapsed gene programs) across Braak LB stages resolved by brain myeloid state. **(a-h)** Activity of the seven Spectra factors that differed between α-synucleinopathy and control donors (FDR < 0.1) and of PKC signaling, shown across Braak LB stages. Lysosomal (enzymes, nucleases, proteases), mitochondrial (quality control, Complex V) and pathogen-recognition programs were lower in α-synucleinopathy, whereas HSR was higher. PKC signaling increased selectively in NUCKS1-high microglia, opposite to the other states. Colored lines show the mean ± s.e.m. of donor-level activity for each microglial state, computed from cells averaged per donor, data type and state. The dotted gray line shows the mean across all states. Activity is the z-scored, cell-level factor score (s.d. units). Braak LB stages: Control, 0; Intermediate, 1-4; Late, 5-6. Statistics are given in the main text and **Supplementary Tables 4-5**.

One factor departed from this pattern only when resolved by state. PKC signaling did not differ between groups across all myeloid cells (−0.07 SD; FDR = 0.15), but was increased in NUCKS1-high microglia (+0.59 SD; *P* = 0.01), whereas it tended to decrease in all other states (−0.11 SD). This divergence between NUCKS1-high microglia and the remaining states was significant (pathology-by-state interaction, +0.70 SD; *P* = 1.9 × 10^−8^; n = 48 donors with NUCKS1-high microglia; **Fig. 4h**). State-resolved trajectories for all factors across Braak LB stages are shown in **Supplementary Fig. 15**. Factor activity differed between brain regions for 17 of 20 factors (FDR < 0.1), whereas the α-synucleinopathy effect differed between regions for only three (GEFs, PI3K signaling kinases and mitochondrial quality control; region-by-pathology FDR < 0.1). For all other main-figure factors, the α-synucleinopathy effect was consistent across regions (interaction FDR ≥ 0.1; **Supplementary Table 5**).

α-synucleinopathy donors thus show lower expression of the lysosomal and mitochondrial machinery that supports degradation and energy supply, together with lower expression of pathogen-recognition modules. Rather than a primed or activated phenotype, this pattern points to reduced sensing and clearance capacity. The concurrent rise in the HSR factor indicates proteostatic stress, consistent with the expansion of HSP90AA1-high microglia (**Fig. 2**) and the differential expression results described above (**Fig. 3**).

### HSP90AA1- and NUCKS1-high microglia are enriched for Parkinson’s disease polygenic risk

To determine whether microglial states differentially capture genetic architecture, we applied single-cell disease relevance scoring (scDRS^32^) using neurodegenerative and neuropsychiatric GWAS summary statistics together with non-neurological traits (rheumatoid arthritis, height) as comparators^33–39^. NUCKS1-high microglia showed significant enrichment for PD polygenic risk (FDR < 0.05; **Fig. 5a**); HSP90AA1-high microglia showed nominal enrichment that did not survive multiple-testing correction. Among neurodegenerative traits, this enrichment was specific to PD: neither population was enriched for AD, ALS or PSP risk, and no other myeloid state showed PD enrichment. Several states were enriched for non-neurological traits (**Fig. 5a**), indicating that myeloid state enrichment is not PD-specific per se. Notably, this within-myeloid signal was not evident in whole-brain single-cell analyses that include all cell types (Hirata *et al.*, in preparation), likely reflecting the dilution of within-lineage heterogeneity by the broader transcriptional variance across cell types—a limitation of disease score approaches applied to heterogeneous datasets.

**Figure 5.**
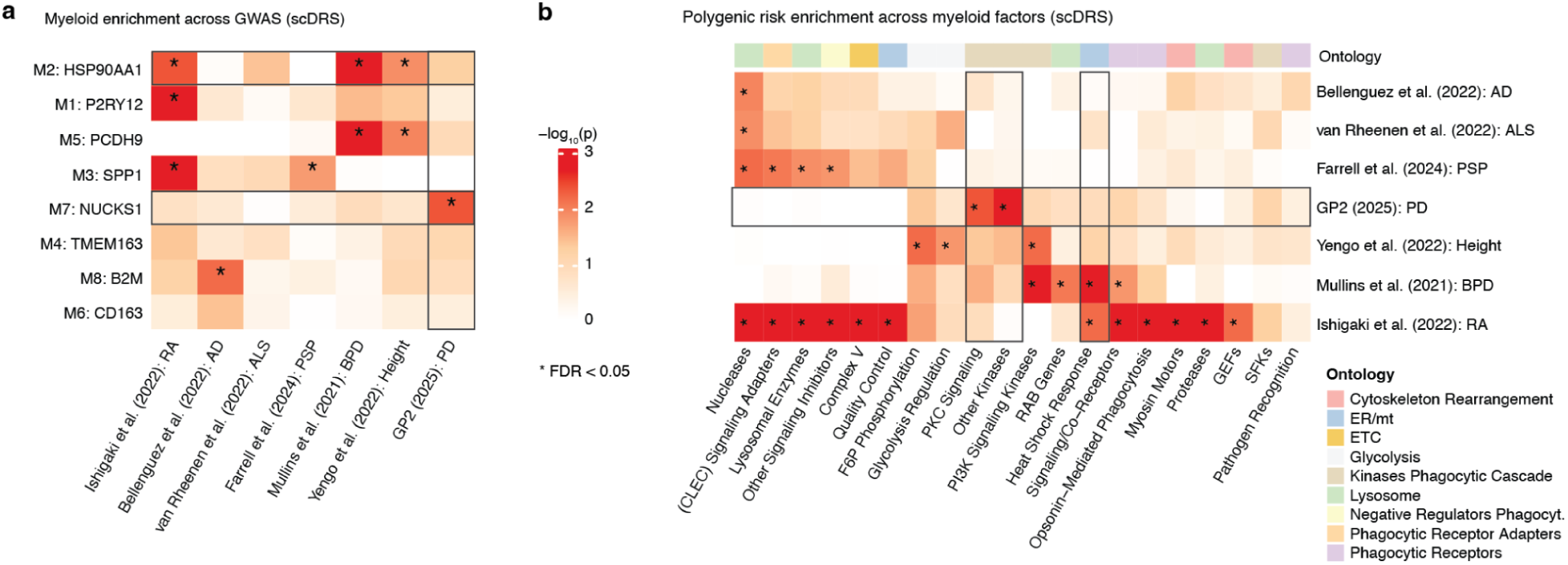
Integration of GWAS polygenic risk with myeloid states and expression-level programs. **(a)** scDRS enrichment of GWAS traits across myeloid states. **(b)** scDRS enrichment of PD GWAS signal across myeloid gene programs. * indicates significance at FDR < 0.05.

We next asked whether specific transcriptional programs mediate genetic risk. Factor-level scDRS revealed that PKC signaling, Other Kinases, and HSR factors were significantly associated with PD risk (FDR < 0.05 for PKC and Other Kinases factors, *P* < 0.05 for HSR factor; **Fig. 5b**). PKC signaling activity was highest in NUCKS1-high microglia (**Fig. 4h**), linking this population’s genetic risk enrichment to a specific functional axis. Examination of PD risk genes revealed that NUCKS1-high microglia expressed elevated levels of *NUCKS1*, *TLR2*, *TBC1D8*, and *SNCA* (**Supplementary Fig. 1c**)—genes involved in DNA damage response^23^, innate immune sensing of α-synuclein^40^, endosomal trafficking^41^, and the pathological substrate itself^42^. HSP90AA1-high microglia instead showed high expression of *BAG3*, a co-chaperone regulating autophagy and proteostasis^43,44^, consistent with this state’s nominal PD risk association operating through a distinct, proteostatic axis. These risk genes loaded highly on the ontological supercategories of the corresponding factors (**Supplementary Fig. 16**), tying each myeloid state’s genetic signal to its implicated functional program.

NUCKS1-high microglia thus emerge as a genetically implicated population in PD: they carry significant polygenic risk enrichment, expand in α-synucleinopathy (**Fig. 2c**), and express *TLR2*, a major receptor for extracellular oligomeric α-synuclein, and the endosomal regulator *TBC1D8* alongside *NUCKS1* (**Supplementary Fig. 1c**). Genetic variation in this state may thus influence PD risk through coupled modulation of innate sensing of α-synuclein and vesicular processing, converging on the PKC signaling program preferentially active in this population.

### The PD-protective P2RY12 eQTL rs3732765 is associated with microglial state composition and lysosomal factor activity

Having identified the transcriptional programs that carry PD polygenic risk, we asked whether individual PD risk variants influence myeloid state composition. We tested the lead variants of 30 microglial *cis*-eQTLs that colocalize with PD GWAS signals from Nalls *et al*. (2019)^45^ or the more recent GP2 meta-analysis^39^ in SingleBrain and MiGA^3,46^, comparing the two opposing genotypes of each variant (e.g. A/A vs G/G) for the abundance of all eight myeloid states across 113 genotyped donors, adjusting for pathology, brain region, and donor covariates with a donor random effect (**Methods**; **Supplementary Table 6**). No test result was significant, the full results for all 30 variants and eight states are reported in **Supplementary Fig. 17** and **Supplementary Tables 6-7**. Because no association was expected to survive correction at this sample size, we focused on a single prespecified hypothesis.

Several features of the *MED12L*/*P2RY12* locus make it the most tractable in our cohort and motivated targeted follow-up of rs3732765. rs3732765 is the top *cis* microglial eQTL for *P2RY12* in MiGA, with the A allele downregulating *P2RY12* expression, and the eQTL colocalizes with the PD GWAS signal at *MED12L*/*P2RY12*^3^. The same A allele confers reduced PD risk in a GWAS meta-analysis (β = −0.06, P = 2.4 × 10⁻¹⁰)^45^.

Because rs3732765 acts as a *cis*-eQTL for *P2RY12*, the defining marker of the P2RY12-high state, we specified the abundance of P2RY12-high microglia as the primary outcome for this variant before testing. Consistent with this hypothesis, A/A donors showed a higher proportion of P2RY12-high microglia than G/G donors across brain regions (logFC = 0.62, *P* = 0.027; 18 A/A vs 57 G/G donors; **Fig. 6a**). The effect was directionally consistent in SN alone but was not significant with only 4 A/A donors (logFC = 0.79, *P* = 0.16; **Supplementary Table 6**). It was weaker under an additive model (*P* = 0.18; **Supplementary Table 7**), suggesting that the shift is most apparent with two copies of the A allele. The protective allele is thus associated with a higher proportion of P2RY12-high microglia, a state that is less abundant at higher Braak LB stages, where HSP90AA1-high cells predominate (**Fig. 2c**). This positions the locus as a candidate link between PD genetic risk and the stage-associated remodeling of microglial states.

**Figure 6.**
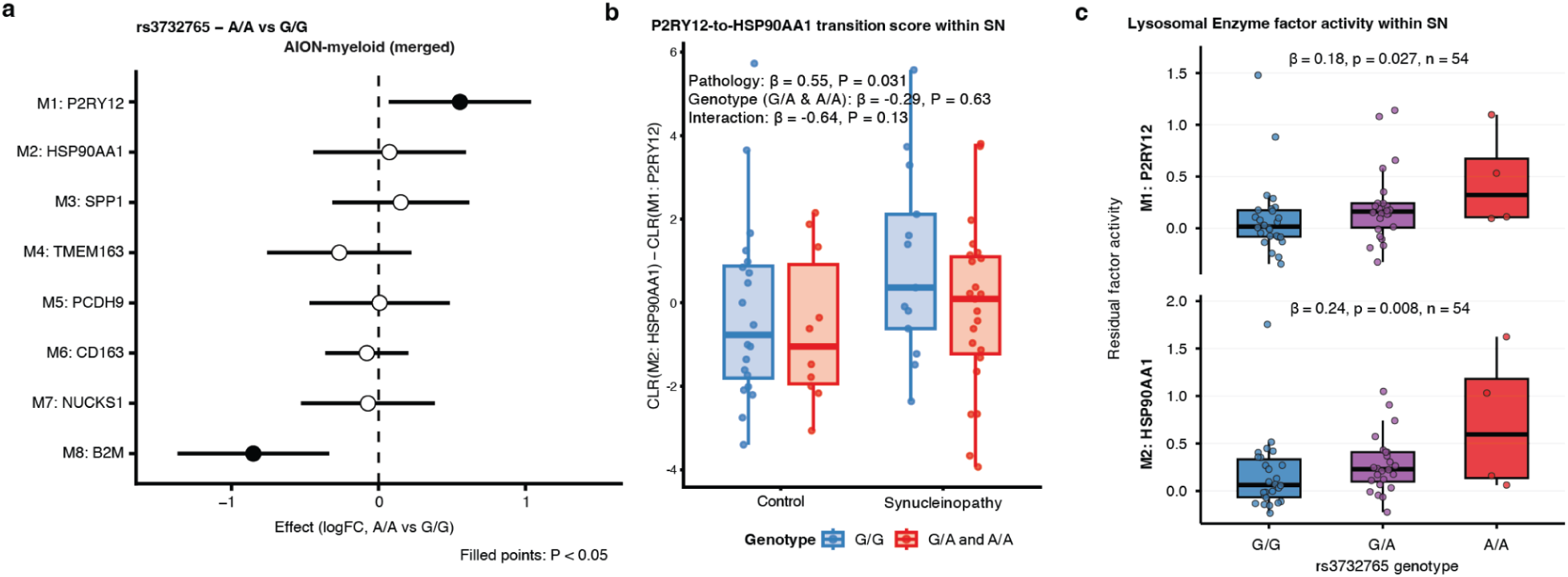
rs3732765 state abundance and factor quantitative trait locus. **(a)** rs3732765 A/A genotype effect on myeloid state abundance. **(b)** Effect of rs3732765-A carrier status (G/A and A/A vs G/G) on the P2RY12-to-HSP90AA1 transition score in SN. **(c)** rs3732765-A allele dosage effect on residual lysosomal enzyme Spectra factor activity within SN.

To examine this remodeling directly, we modeled the CLR-based transition score from P2RY12-high to HSP90AA1-high microglia, which increased with pathology group (β = 0.55, *P* = 0.031; **Fig. 6b**). The genotype-by-pathology interaction estimate was negative, consistent with a smaller increase in A-allele carriers, but did not reach significance (β = −0.64, *P* = 0.13).

The association with genotype was more evident at the level of lysosomal enzyme factor activity, which is lower in microglia from donors with α-synucleinopathy (**Fig. 4c**). Residual factor activity, adjusted for disease status and donor covariates, increased with A-allele dosage in both P2RY12-high (β = 0.18, *P* = 0.027, n = 54) and HSP90AA1-high microglia (β = 0.24, *P* = 0.008, n = 54; **Fig. 6c**), with the larger effect in the stress-associated state. Together, these data nominate rs3732765-A as a microglial *cis*-regulatory variant that lowers *P2RY12* expression (MiGA^3^), associated here with a higher proportion of P2RY12-high microglia, and higher lysosomal factor activity in both sensome and proteostatic stress-associated states.

## Discussion

We present AION-myeloid, a myeloid-enriched single-cell atlas of the human SN and connected brain regions which, integrated with AMP PD, comprises 298,592 myeloid profiles (**Fig. 1a**; **Table 1**). Myeloid enrichment recovered more SN myeloid profiles than prior PD SN studies combined and resolved eight states that replicated across cohorts and isolation methods (**Fig. 1c,d**; **Supplementary Fig. 2i-k**). Three findings emerge: α-synucleinopathy is associated with a shift from sensome toward proteostatic-stress microglia; this shift is accompanied by lower expression of clearance and sensing programs rather than a classical activation signature; and PD genetic risk maps onto specific states and programs within this landscape.

The most consistent compositional change was a lower proportion of P2RY12-high sensome microglia and a higher proportion of HSP90AA1-high microglia, with larger effects at late Braak LB stages (**Fig. 2c**; **Supplementary Fig. 8**). The two states were inversely correlated in α-synucleinopathy donors but not in controls (**Fig. 2d,e**). At the protein level, HSP60^+^ microglia increased with SN neuron loss in parallel with the sequencing-based HSP90AA1-high fraction (**Fig. 2f-i**; **Supplementary Fig. 9**). If this reflects state conversion, sensome microglia would lose their surveillance program and enter a proteostatic-stress state under α-synuclein burden, rather than being replaced by a distinct population. This interpretation fits the reduced sensing and clearance programs described below. However, because the design is cross-sectional and the data compositional, this anti-correlation may reflect state conversion, selective loss or expansion, or differential turnover; distinguishing these will require trajectory, lineage-tracing or perturbation data. Independently of pathology, HSP90AA1-high microglia were more abundant in the DMNX and GPI than in cortex (**Supplementary Fig. 10**), raising the possibility that regional myeloid composition relates to regional susceptibility to Lewy pathology, a relationship our data cannot resolve. Sex and age effects were modest and did not survive multiple-testing correction (**Supplementary Fig. 11a**).

At the transcriptional level, α-synucleinopathy was associated with higher expression of heat-shock-response (HSR) and glycolytic genes, whose activities were tightly coupled across donors, and lower expression of lysosomal, electron-transport and interferon genes (**Fig. 3a-e**). Gene-program analysis gave a concordant picture: programs for lysosomal enzymes, nucleases, proteases, mitochondrial quality control, Complex V and pathogen recognition were lower in disease, and only the HSR program was higher (**Fig. 4a-g**). Together, these data describe microglia with reduced degradative, bioenergetic and sensing capacity under proteostatic stress, rather than a primed or pro-inflammatory phenotype. In terms of the question posed at the outset, these data argue against a preserved response and are most consistent with an exhausted rather than a hyperactivated, maladaptive state, although exhaustion and maladaptation are not mutually exclusive. Furthermore, whether this reduced capacity is a consequence of accumulated aggregate burden or a pre-existing limitation that contributes to impaired α-synuclein clearance cannot be determined from post-mortem tissue. The same directional pattern was present in peripheral monocytes from people with PD^29^ (**Supplementary Fig. 14**). Because circulating monocytes have little direct contact with parenchymal aggregates, this concordance is compatible with a systemic or intrinsic component of myeloid dysfunction in PD, although the moderate pathway-level correlation and the differences between cohorts and assays warrant caution.

NUCKS1-high microglia were the only state significantly enriched for PD polygenic risk (**Fig. 5a**). Because this state is rare (0.5–1.3% of myeloid cells), we characterized it before interpreting its disease association: it expressed canonical microglial markers (**Supplementary Fig. 1b**), was recovered across cohorts and modalities (**Supplementary Fig. 2e,i-k**), was not enriched for apoptosis markers (**Supplementary Fig. 5**), and was largely unrelated to technical covariates (**Supplementary Fig. 6**). NUCKS1-high microglia were more abundant in late-stage α-synucleinopathy (**Fig. 2c**), were the only state enriched for an S-phase signature (**Supplementary Fig. 7**), and showed higher PKC-signaling activity in disease, opposite to all other states (**Fig. 4h**), a program itself associated with PD risk (**Fig. 5b**). Their risk enrichment is carried by elevated expression of PD risk genes including *NUCKS1*, *TLR2*, *TBC1D8*, and *SNCA* (**Supplementary Figs. 1c**, **1****6**); the scDRS signal and the risk-gene expression are therefore two views of the same observation rather than independent evidence. The S-phase signature raises the possibility that NUCKS1-high cells represent a proliferative compartment, consistent with local microglial renewal in affected regions, but this remains to be tested. Abundance changes were also regionally heterogeneous, higher in SN but lower in the DMNX and GPI (**Supplementary Fig. 10b**). Whether NUCKS1-high microglia are protective, pathogenic or bystanders remains open.

At the single-variant level, the PD-protective rs3732765-A allele, the lead microglial cis-eQTL for *P2RY12* in MiGA, which lowers *P2RY12* expression and colocalizes with the *MED12L*/*P2RY12* GWAS signal^3^, was associated with a higher proportion of P2RY12-high microglia (**Fig. 6a**) and with higher lysosomal-enzyme program activity in P2RY12-high and HSP90AA1-high microglia in the SN (**Fig. 6c**). The genotype-by-pathology interaction on the P2RY12-high-to-HSP90AA1-high balance was directionally consistent with attenuation but not significant (**Fig. 6b**). These associations are nominal, rest on few A/A donors, and were tested as a single prespecified hypothesis within a panel reported in full (**Supplementary Fig. 17**; **Supplementary Tables 6-7**); they require replication in larger genotyped cohorts and functional validation.

This study carries limitations. First, the design is cross-sectional, so associations with Braak LB stage cannot establish temporal order or causality. Second, abundance estimates are compositional, so the lower abundance of one state is partly the counterpart of higher abundance of others. Third, we combined FANS-sorted nuclei, MACS-enriched cells and AMP PD nuclei; although dataset, data type and brain region were modeled as covariates, isolation method is partly confounded with brain region, and tissue dissociation can induce *ex vivo* stress signatures, which may particularly affect rare states and cross-regional comparisons. Fourth, the genetic analyses have limited power and scope: the rs3732765 findings are nominal, and scDRS enrichment neither establishes causality nor is exclusive to PD, as other states were also enriched for non-neurological traits (**Fig. 5a**). Fifth, the IHC validation included 12 donors with few controls. Finally, we did not compare α-synucleinopathy with other proteinopathies, so whether these patterns are specific to α-synuclein remains to be determined.

Together, these findings describe a structured remodeling of human brain myeloid states in α-synucleinopathy, from sensome toward proteostatic-stress microglia with reduced clearance and sensing programs, and connect PD genetic risk to a rare NUCKS1-high state and to the *P2RY12* locus. These states and programs are candidates for mechanistic testing in human cellular models.

## Methods

### Human tissue donors

De-identified case and control postmortem brain tissues were obtained from two sources. Fresh-frozen and formalin-fixed paraffin-embedded (FFPE) tissue from frontal cortex, occipital cortex, and midbrain at the level of the red nucleus was provided by the Mount Sinai Neuropathology Brain Bank & Research CoRE (RRID:SCR_027565). Fresh-frozen tissue from frontal cortex and amygdala was provided by the Netherlands Brain Bank (NBB). NBB postmortem donors were stratified by rs3732765-A genotype. For single-nucleus assays, 40 donors were procured by the NBB, with 51 respective tissues across two regions (11 amygdala, 40 cortex). 58 donors (all substantia nigra) were procured by the NPBB. For single-cell RNA-seq, 32 donors were procured by the NPBB, with 52 respective tissues across three regions (19 cortex, 3 occipital, 30 substantia nigra). One substantia nigra control sample was excluded after QC, leaving 97 donors (108 samples) for single-nucleus analyses. In total, 113 unique donors were included post-filtering with a mean age of 76.2 years (range 18 to ≥90 years), of which 40 donors were female. Detailed information for each sample is provided in **Supplementary Table 3**.

### Isolation of nuclei from brain cells for RNA and multiome assay

Nuclei were isolated using the S2 Genomics Singulator 100 (two to four tissue samples per cartridge, each from a different individual) in combination with a previously described Tween-20 lysis buffer^47,48^. Substantia nigra was dissected from the surrounding midbrain parenchyma prior to nuclei isolation. We used 20-100mg of tissues per donor (same amounts for each donor per run), varying by the brain region processed. All buffers were ice cold and supplemented with 1:200 RNAse inhibitor (RRI; Takara). All centrifugation steps were performed in a swinging bucket rotor centrifuge at 600 × g for 10 minutes at 4°C. Briefly, tissues were automatically dissociated in 3mL tween lysis buffer with DTT (1:1000, Sigma Aldrich) and rinsed with S2 Genomics Nuclei Storage Reagent. The nuclei suspension was spun down with max brake, resuspended in diluted Debris Removal Solution (S2 Genomics), and spun down again with slow brake. The pelleted nuclei were then resuspended in 300 µL blocking buffer (5% Gibco BSA), filtered through a 30 µm mesh, and incubated for one hour with SOX10-AF647 (1:1000; abcam, Cat. #ab270151) and NeuN-PE (1:1000; Sigma Aldrich, Cat. #FCMAB317PE) antibodies. Prior to fluorescence-activated nuclei sorting (FANS), volumes were brought up to 1.5 mL with blocking buffer and stained with DAPI (Sigma Aldrich, Cat. #MFCD00012681). Nuclei were then FAN-sorted in blocking buffer coated 5mL tubes using a SOX10-/NeuN+ and SOX10-/NeuN-gating strategy (oligodendrocyte depletion) previously described^49^.

Sorted nuclei fractions were spun down, counted manually using a SYBR Green I nucleic acid gel stain (Cat. #50513), and resuspended in a loading buffer (1% Gibco BSA) volume appropriate to reach concentrations of 1500 nuclei per µL (10x Genomics 3’ v3.1 RNA chemistry) or 3000-5000 nuclei per µL (10x Genomics Multiome ATAC + RNA chemistry; multiome data not analyzed here). For RNA, targeted recovery was 30,000 nuclei. For multiome, targeted recovery was 10,000 nuclei. GEMs and cDNA libraries were prepared according to protocol (10x Genomics). All libraries were sequenced at Azenta using the NovaSeq 6000 platform (Illumina) at a depth of 30,000 reads per nucleus.

### Human postmortem brain dissociation and CD11b+ myeloid-cell enrichment

All centrifugation steps were performed at 4°C, and all tissue-handling steps were carried out on ice unless otherwise indicated. We optimized a previously established protocol for microglia isolation from mouse brain for use with fresh postmortem human brain tissue. This approach was designed to minimize technical and biological artifacts introduced by tissue dissociation and cell isolation, as previously described^29^.

Fresh dissociation buffer, hereafter referred to as PBS+, was prepared on the day of isolation and consisted of PBS supplemented with 0.5% BSA, 25 mM HEPES, and 2 mM EDTA. Percoll was pH-adjusted to 7.3-7.4 and kept at 4°C. Isotonic Percoll was prepared immediately before use by mixing nine parts pH-adjusted Percoll with one part 10X PBS.

Human brain tissue was transferred to a pre-chilled Petri dish containing ice-cold PBS+ and minced into small fragments using sterile scalpels. The minced tissue was transferred into a pre-chilled 15 mL Dounce homogenizer containing 12 mL ice-cold PBS+ and mechanically dissociated using a loose pestle. Homogenization was performed with gentle up-and-down strokes while rotating the pestle, until no large tissue fragments remained, while avoiding excessive mechanical stress. The resulting cell suspension was transferred to a 50 mL conical tube and centrifuged at 450 × g for 10 minutes.

After centrifugation, the supernatant was removed and the pellet was resuspended in PBS+ to a final volume of 12 mL. Four milliliters of isotonic Percoll were added and mixed thoroughly. Sixteen milliliters of PBS+ were then carefully overlaid onto the Percoll-containing suspension by slowly pipetting along the wall of the tilted tube to preserve phase separation. Samples were centrifuged at 3000 × g for 10 minutes. The upper phase and myelin-containing interface were carefully aspirated, leaving approximately 5 mL above the pellet to minimize loss of low-density cells. Ten milliliters of ice-cold PBS+ were then gently added along the wall of the tube without directly disturbing the pellet. Tubes were gently tilted to mix the remaining Percoll with the wash buffer, and cells were pelleted by centrifugation at 500 × g for 8 minutes. The resulting pellet contained the total brain cell fraction after myelin removal.

For enrichment of CD11b+ myeloid cells, the total brain cell pellet was resuspended in 360 µL MACS buffer consisting of PBS supplemented with 1% BSA. Thirty-five microliters of CD11b nanobeads were added, and cells were incubated for 15 minutes at 4°C. Cells were then washed with 2 mL MACS buffer and pelleted at 400 × g for 8 minutes. The pellet was resuspended in 500 µL MACS buffer and applied to LS columns mounted on a Miltenyi quadroMACS magnet. Columns were washed three times with 3 mL MACS buffer, and CD11b+ cells were eluted with 3.5 mL MACS buffer for downstream analyses.

### DNA isolation and genotyping

Genomic DNA was extracted from the frontal cortex, amygdala, or substantia nigra and genotyped using the Illumina Infinium Global Screening Assay as previously described^3^.

### Computational preprocessing

Alignment of sequencing reads from each multiplexed sample batch was done using the CellRanger suite (v9.0.1) against the hg38 reference genome. We leveraged the CellBender tool (v0.3.0)^50^ to remove ambient RNA and empty droplets from each 10x run. We used cellSNP-lite (v1.2.2)^51^ to detect SNPs directly from our snRNA-seq data. Subsequently, vireoSNP (v0.5.8)^52^ was applied to demultiplex pooled snRNA-seq samples by integrating observed allele counts with matched donor genotypes, enabling barcode donor assignment and genetic doublet detection. Genotype concordance was verified via QTLtools-mbv (v1.3.1)^53^. As a first-pass QC, nuclei from each 10x snRNA-seq run were removed if they had fewer than 15 genes (permissive threshold, as empty droplets and ambient RNA were already removed with CellBender and nuclei were FAN-sorted), more than 15% mitochondrial content, or genotype discordance (nuclei with unassigned donors or with a doublet log-likelihood > 3.84 were removed). For scRNA-seq, the cells from each 10x run were removed if they had fewer than 200 genes or more than 20% mitochondrial reads. Scanpy’s (v1.11.1) built-in scrublet^54,55^ function was applied to each 10x sn/scRNA-seq run to remove expression doublets.

### sn/scRNA-seq processing and analysis pipeline

A total of 452,290 cells and 423,445 nuclei passed the initial QC steps and were merged into two separate datasets for downstream analysis. Normalization and clustering were performed using Scanpy and Pegasus (v1.10.2)^56^. Counts were normalized to library size and log-transformed. The top 2,000 variable genes were selected with the *pg.highly_variable_features()* function in Pegasus. Technical effects from library size and number of nuclei or cells were regressed out using the *pg.regress_out(*) function. Batch effects from each different 10x run were addressed with the Harmony algorithm via the *pg.run_harmony()* function.

Dimensionality reduction was performed by first applying Principal Component Analysis (PCA) on the variable genes. We constructed a k-nearest neighbors (k-NN) graph (k=30) and then applied Uniform Manifold Approximation and Projection (UMAP; 30 nearest neighbors, min_dist=0.3, spread=1.0), using the top principal components (PCs) that cumulatively captured > 95% of the variance explained. Clusters were identified using the Leiden algorithm. These steps were executed using the Pegasus functions *pg.highly_variable_features()*, *pg.pca()*, *pg.neighbors()*, *pg.umap()*, and *pg.leiden()*.

Each barcode was annotated with a class-level cellular identity by annotating each cluster through visual inspection of cell-type markers (markers are in **Supplementary Fig. 1**). Myeloid nuclei and cells were then in *silico* selected and underwent a re-analysis with the same pipeline described above. Non-myeloid nuclei or cells were filtered out. Myeloid nuclei (AION cohort) were used as a discovery cohort and subclustering was performed to identify distinct myeloid states. Clusters were manually annotated into myeloid states based on top marker gene expression and labeled by a representative marker (e.g., P2RY12-high), set in roman type to denote cell populations rather than gene expression.

### Integrated myeloid dataset embedding and visualization

To create an integrated myeloid embedding of all microglial cells and nuclei, we joined our AION-myeloid dataset with that of AMP PD^6^ and SingleBrain (consisting of Fujita *et al.*^57^, Bryois *et al.*^58^, Gabitto *et al.*^59^, & Mathys *et al.*^60^). Expression matrices were subsetted to 15,642 genes common between all cohorts, and features selected using the top 3000 most variable genes of the in-house cells and nuclei. CONCORD^14^ was then used with the individual IDs as the CONCORD domain to transfer the 8 myeloid labels to the SingleBrain collection, using default parameters. AION-myeloid and AMP PD data was then extracted from the joint object. A 2D UMAP was then created in the joint CONCORD embedding space for subsequent visualization of the myeloid clustering.

### Neuropathological staging

Owing to the small number of early- and medium-stage donors, early and medium Braak LB stages were combined into a single intermediate group for abundance and transcriptional analyses; contrasts were therefore intermediate-vs-control and late-vs-control.

### Differential abundance analysis

Differential abundance analysis was performed using crumblr (v0.99.19)^26^ as previously described^6^. Briefly, the *aggregateToPseudoBulk()* function from the dreamlet package (v1.4.1)^61^ was applied to derive a pseudobulk object for each cohort that contained all state-level count assays, including normalized cell counts. A crumblr object was generated with the *crumblr()* and *cellCounts()* functions. To test for abundance changes across different conditions, we applied the following formulae with the *dream()* function from the variancePartition package^27^:

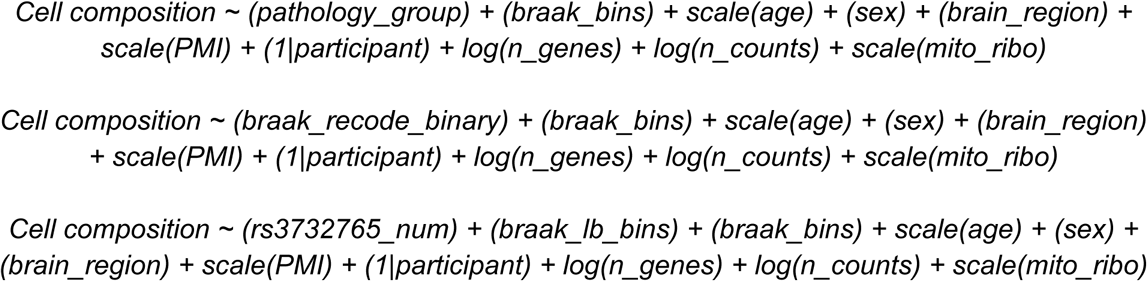

Here *pathology_group* encoded pathology status as a binary variable and *braak_recode_binary* encoded a temporary Braak LB stage binary variable where each pathological stage was tested against controls (0: Control, 1-4: Intermediate, 5-6: Late). *Braak_bins* represented Braak stages (0: Control, < 4: less than 4, ≥ 4: ADNC). *Participant* was used as the donor ID. *brain_region* contained DMNX, substantia nigra, amygdala, GPI, and cortex. *Participant* was modeled as a random effect, while all other variables were modeled as a fixed effect.

### Differential gene expression analysis

We utilized dream for differential expression analysis applied on our pseudobulk objects. To test for expression changes across pathology and isolation methods (nuclei vs cells), we applied the following respective formulae with the *dream()* function from the variancePartition package^27^.

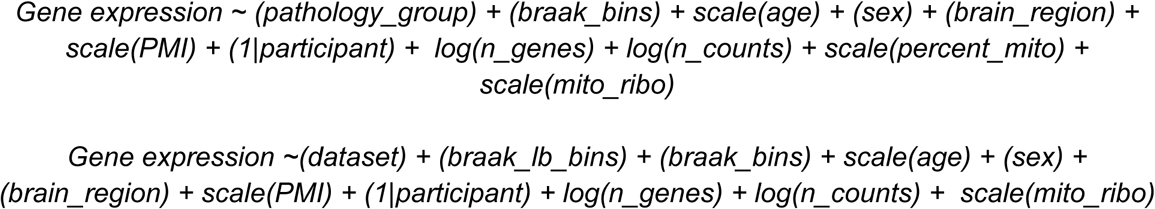

### Over-representation analysis (ORA)

Using clusterProfiler (v4.14.6) and a custom R function^3^ we applied Gene Ontology (GO) enrichment and ORA on top cell-state markers, global and cell-state-specific differentially expressed genes (DEGs). Briefly, we performed a pairwise enrichment analysis across two collections of gene sets. For each pair, we removed missing values and computed the number of overlapping genes. We then constructed a 2-by-2 contingency table representing shared and non-shared genes relative to a specified background of genes and applied a one-sided Fisher’s exact test to test for over-representation of the overlap. For each comparison, we recorded the gene set identifiers, sizes, overlap count, odds ratio, nominal *P*-value, and the list of overlapping genes. Multiple testing correction was performed using the Benjamini-Hochberg method. Each top marker or DEGs set was tested against GO:BP terms or manually curated gene sets implicated in Parkinson’s disease^29^.

### Chromogenic Immunohistochemistry (IHC)

The paraffin-embedded sections of human frontal cortex and midbrain (5µm thickness) were pretreated using Cell Conditioning CC1 antigen retrieval buffer (Tris/Borate/EDTA buffer, pH 8.0–8.5, 950-224, Roche Diagnostics), and incubated with primary antibody against the mitochondrial marker HSP60 [EPR18245-93] 1:5000 (abcam #ab190828) and secondary antibody Multimer HRP OmniMAP-Anti Rabbit (760-4311, Roche Diagnostics). The detection was performed using the Detection Kit ChromoMAP DAB Kit (760-159, Roche Diagnostics). Hematoxylin and Bluing reagent (760-2021, 760-2037, Roche Diagnostics) were used as a nuclear counterstain.

### Chromogenic Immunohistochemistry Multiplex (IHC)

The paraffin-embedded sections of frontal cortex and midbrain (5µm thickness) were pretreated using Cell Conditioning CC1 antigen retrieval buffer (Tris/Borate/EDTA buffer, pH 8.0–8.5, 950-224, Roche Diagnostics), and incubated in rounds of staining with primary antibodies against the mitochondrial marker HSP60 [EPR18245-93] 1:5000 (abcam #ab190828), microglial marker IBA1 1:500 (Thermo Fisher Scientific #PA5-27436), astrocyte marker ALDH1L1[3E9] 1:700 (abcam #ab56777) and secondary antibody Multimer HRP OmniMAP-Anti Mouse and Multimer HRP OmniMap-Anti Rabbit (760-4310 & 760-4311, Roche Diagnostics). The detection was performed using DISCOVERY Purple HRP Kit (ALDH1L1), DISCOVERY Green HRP Kit (HSP60), and DISCOVERY Yellow HRP Kit (IBA1) (Cat # 760-229, 760-271, 760-250, Roche Diagnostics). Sodium citrate antigen retrieval (pH 6.0) was used between rounds to remove the antibody from the previous round and avoid cross-reactivity. Hematoxylin and Bluing reagent (760-2021, 760-2037, Roche Diagnostics) were used as a nuclear counterstain. Whole-slide digital images were acquired at 40X magnification using the Aperio GT450 high-resolution slide scanner (Leica Biosystems).

### Whole-slide image processing and cell detection

Triple-chromogen brightfield whole-slide images (IBA1, yellow; HSP60, green; ALDH1L1, purple; scanned at 40X, 0.26 µm/px) were analyzed in QuPath v0.7.0. Each slide was set to the *Brightfield (Other)* image type and color-deconvolved using per-slide stain vectors: unit optical-density (OD) vectors were derived from RGB values sampled in single-positive regions for each chromogen, with a per-slide background (white) reference, and a reference vector set (donor D079) was applied to slides for which a dedicated, spectrally clean calibration could not be obtained. The substantia nigra pars compacta was annotated manually on each slide. Within this region, cells were detected with the QuPath watershed cell-detection routine operating on the optical-density sum (requested pixel size 0.5 µm, background radius 8 µm, Gaussian σ 3 µm, intensity threshold 0.10 OD, minimum/maximum area 20/1200 µm^2^, cell expansion 4 µm); watershed post-processing (object splitting) was disabled so that ramified microglia were retained as single objects rather than fragmented. For every detected cell, mean, maximum and standard-deviation OD were recorded for each deconvolved channel, together with morphological features (area, circularity, calipers, eccentricity).

#### Cell classification

Cells were classified per slide to accommodate between-slide staining differences. Lineage-marker positivity was determined by fitting a two-component Gaussian mixture model (expectation-maximization) to the per-cell mean OD of IBA1 and of ALDH1L1 and taking the density crossing point of the two components as the positivity threshold.

Neuromelanin-laden neurons—an endogenous broadband pigment that three-chromogen deconvolution cannot spectrally separate—were identified with a QuPath object classifier (random-trees) trained on operator-labeled pigmented-neuron and non-pigment examples drawn from multiple slides and applied across the cohort; because neuromelanin and glial identity are mutually exclusive, classifier-positive cells were assigned to a Neuromelanin class and excluded from glial analysis. Cells positive for both IBA1 and ALDH1L1 (a biologically implausible microglia-astrocyte double identity) were flagged as ambiguous and excluded. HSP60 positivity, used to define proteostatically burdened microglia and astrocytes, was assessed on the per-cell maximum HSP60 OD rather than the mean, because mitochondrial HSP60 is punctate and broadly expressed, rendering the mean unimodal and unstable across slides; a two-component Gaussian mixture was fit to the maximum-OD distribution of the lineage-classified glia and the threshold was placed a fixed fraction (0.35) of the way from the component crossing toward the baseline mode. Cells were thereby assigned to microglia (IBA1^+^), proteostatic microglia (IBA1^+^/HSP60^+^), astrocytes (ALDH1L1^+^), proteostatic astrocytes (ALDH1L1^+^/HSP60^+^), HSP60^+^-only, negative, neuromelanin^+^ neurons, or ambiguous. Detection and classification were applied identically to all 12 slides by batch scripting the QuPath project, and per-cell measurements were exported for downstream analysis in R v4.4.3.

#### Cross-modality comparison

In donors with both SN immunohistochemistry (IHC) and SN sn/scRNA-seq data (n = 11), we calculated the proportion of proteostatically burdened microglia as IBA1⁺HSP60⁺ / IBA1⁺ cells (IHC) and as M2: HSP90AA1-high / all myeloid cells (sn/scRNA-seq). One library was used per donor; for donors profiled by both methods, the library with more myeloid cells was used. Both measurements were analyzed in a single quasibinomial regression of cell counts on SN neuron-loss grade (0-3; 0 = None, 1 = Mild, 2 = Moderate, 3 = Severe), measurement method (IHC or sn/scRNA-seq), and whether this relationship differed between methods, with each donor counted as one unit. We report the modeled percentage of HSP90AA1-high microglia at no and at severe SN neuron loss for each method, and tested whether the rise with SN neuron loss differed between them.

### Spectra analysis

To discover biologically informed gene programs that were dysregulated across disease states, we employed a latent factor approach called Spectra (v0.2.0)^31^. Spectra integrates prior knowledge by building gene-gene similarity graphs from pathway annotations, constructing separate graphs for each cell-type (or cell-state) and a global graph. It jointly learns cell- and gene-specific parameters that represent distributions over gene expression programs, encouraging genes connected in the graph to have similar representations. We annotated each latent factor using our manually curated gene sets and then collapsed the factors into a non-redundant set of gene programs based on semantic similarity.

#### Factor scoring

For each gene program, cell-level activity was computed as the mean score across its constituent factors, winsorized at the 99th percentile of absolute values, transformed with a sign-preserving log1p (sign(*x*)·log(1 + |*x*|)) and z-scored across all cells. Cells with ambiguous or missing donor assignment were excluded.

#### Pathology and Braak LB association

Cell scores were averaged per donor and data type (fresh cells or nuclei; AMP PD nuclei were treated as nuclei), retaining units with ≥20 cells. For each program we fitted a linear mixed model, *score ∼ pathology group + data_type + age + sex + PMI + (1 | participant)*, using lme4 and lmerTest with Satterthwaite degrees of freedom; a linear model was used when no donor contributed more than one unit. *P* values were adjusted across the 20 programs using the Benjamini-Hochberg procedure, and programs with FDR < 0.1 were considered significant. Of 193 donors, 3 were excluded for having fewer than 20 myeloid cells in every data-type unit and 2 for missing PMI, leaving 188 unique donors. Braak LB stages were binned as Control (0), Intermediate (1-4) and Late (5-6) and modeled in the same way, testing each stage against Control and a per-stage linear trend.

#### State-resolved analysis

To test programs within microglial states, scores were averaged per donor, data type and state (≥10 cells) and modeled as *score ∼ pathology group + (1 | participant)*. Because rare states are not captured in every donor, fixed covariates were omitted at this level to avoid excluding donors with sparse representation. Whether the pathology effect in a given state differed from that in the remaining states was assessed with a pathology-by-state interaction term in a model including all states and a donor random intercept.

#### Brain-region effects

Region dependence was assessed on scores averaged per donor, data type and brain region (≥20 cells), using *score ∼ brain_region + pathology_group + data_type + (1 | participant)*, with an additional region-by-pathology term to test whether the pathology effect differed between regions. Omnibus *F*-tests were adjusted across programs using the Benjamini-Hochberg procedure (**Supplementary Table 5**).

### Single-cell disease relevance scoring (scDRS)

To calculate combined expression of disease-associated genes identified through GWAS summary statistics, we employed scDRS (v1.0.2)^32^ and MAGMA^62^. The *scdrs compute-score* command was run with default settings, except that 500 control scores were computed. For downstream analysis, we aggregated cell-scores across state-, Spectra factor-, and pathology levels using the *scdrs perform-downstream* command with default settings.

### Selection of PD risk variants and genotype-state association testing

Candidate variants were drawn from the eQTL-GWAS colocalization results of SingleBrain^46^, which include colocalization of single-nucleus microglial eQTLs and of a reanalysis of MiGA microglial eQTLs^3^ (primary and conditionally independent signals) with PD GWAS summary statistics from Nalls *et al*. (2019) and GP2 (2025; proxy and clinical cases, and clinically defined cases only)^39,45^. We retained locus-gene pairs with a posterior probability of a shared causal variant (PP4) > 0.8 in either microglial eQTL set and, for each, selected the lead eQTL variant, yielding 49 unique variants. 30 of these 49 unique variants passed imputation quality and a minimum number of homozygous donors per genotype group and were tested. Because the additive model does not require homozygous donors in both groups, 43 microglial eQTL variants and 136 GP2 (2025)^39^ lead variants were tested under that model (**Supplementary Table 7**).

Donors were genotyped on the Illumina Global Screening Array (GSA). Genotypes were imputed using the TOPMed r3 reference panel on the NIH BioData Catalyst TOPMed Imputation Server^63–65^. Imputed variants were filtered to an imputation quality of R^2^ ≥ 0.3 and were taken as hard calls (hg38) coded as alternate-allele dosage. Ancestry information was available for all 113 donors in the AION cohort, comprising 82 European (EUR; 72.6%), 18 African (AFR; 15.9%), 8 Admixed American (AMR; 7.1%), 3 East Asian (EAS; 2.7%), and 2 South Asian (SAS; 1.8%) individuals. Genetic principal components (gPCs) were computed with PLINK v2.0^66^ (--pca) across all 113 donors on 363,963 variants with minor allele frequency ≥ 5%, retained after linkage disequilibrium pruning (--indep-pairwise 50 5 0.2; 50-variant windows, step size 5, r^2^ < 0.2). The first two gPCs were included as covariates in all genotype-based models to account for population structure.

For each variant, donors homozygous for the alternate allele were compared with donors homozygous for the reference allele, excluding heterozygotes; an additive model across all three genotypes was fitted as a supplementary analysis. Myeloid state abundances were transformed to centered log-ratios with crumblr^26^ and modeled with dream (variancePartition^27^), adjusting for gPCs 1-2, Braak stage, Braak LB stage, age, sex, brain region, post-mortem interval, log-transformed gene and UMI counts, mitochondrial read fraction, and mitochondrial and ribosomal gene counts, with a random intercept for donor. Analyses were run across all brain regions and in SN alone; variants with fewer than two donors in either genotype group were not tested. P values were corrected using the Benjamini-Hochberg procedure across all variant-state tests within each variant set and region.

### Statistical analysis

Analyses were performed in R (v4.4.3) and Python unless otherwise stated. Unless otherwise noted, P values were corrected for multiple testing using the Benjamini-Hochberg procedure. Significance thresholds were FDR < 0.05 for differential gene expression and scDRS, and FDR < 0.1 for differential abundance and Spectra factor analyses. Nominal P values are reported for the hypothesis-driven curated-pathway ORA of state-specific DEGs, for the prespecified rs3732765 analysis, and for sex and age associations, which did not survive correction. In figures, asterisks denote FDR thresholds and dots or filled points denote nominal P < 0.05, as specified in each legend.

## Data Availability

All summary statistics are provided in the Supplementary Tables. Raw single-cell RNA-sequencing data and processed count matrices will be made available through CELLxGENE upon publication.

AMP PD data can be accessed through CELLxGENE: https://cellxgene.cziscience.com/collections/d5d0df8f-4eee-49d8-a221-a288f50a1590.

## Code Availability

The code used for the primary analysis is available on GitHub at: https://github.com/RajLabMSSM/AION-myeloid. Any additional code used for analysis is available upon request from the corresponding author.

## Acknowledgement

We thank the study participants for their generous gifts of brain donation. The authors thank the teams of the Netherlands Brain Bank and the Mount Sinai Neuropathology Brain Bank and Research CoRE for their services. We thank Stella Dracheva and her team for training in nuclei isolation, Calvin Jan, Subhabrata Sanyal and Madhurima Benekareddy (Calico Life Sciences), and Jack Humphrey for helpful discussions and support. We thank Brian Fulton-Howard for performing genotype imputation and quality control. T.R. and J.B. received funding from Calico Life Sciences. T.R. is supported by the National Institutes of Health (NIH; grants NIA U01-AG068880, NIA R21-AG063130, NIA R01-AG054005, NIA RF1-AG065926, NIA R01-AG065926, NIA R56-AG088669, NIA R21-AG091272, NIA P30-AG066514, NINDS U54-NS123743 and NINDS R01-NS116006). This work was supported in part by the computational resources and staff expertise provided by Scientific Computing at the Icahn School of Medicine at Mount Sinai and by the Clinical and Translational Science Awards grant (UL1TR004419) from the National Center for Advancing Translational Sciences. This research was supported by the Office of Research Infrastructure of the NIH (awards S10OD026880 and S10OD030463).

Data used in the preparation of this article were obtained from the Accelerating Medicines Partnership® (AMP®) Parkinson’s Disease (AMP PD) and Parkinson’s Disease & Related Disorders (AMP PDRD) Knowledge Platform. For up-to-date information on the study, visit https://www.amp-pdrd.org. The AMP® PD program is a public-private partnership managed by the Foundation for the National Institutes of Health and funded by the National Institute of Neurological Disorders and Stroke (NINDS) in partnership with the Food and Drug Administration (FDA), National Institute on Aging (NIA), Aligning Science Across Parkinson’s (ASAP) initiative; Celgene Corporation, a subsidiary of Bristol-Myers Squibb Company; GlaxoSmithKline plc (GSK); The Michael J. Fox Foundation for Parkinson’s Research (MJFF); AbbVie Inc.; Pfizer Inc.; Sanofi US Services Inc.; and Verily Life Sciences LLC. ACCELERATING MEDICINES PARTNERSHIP and AMP are registered service marks of the U.S. Department of Health and Human Services.

The funders had no role in the design and conduct of the study; the collection, management, analysis and interpretation of the data; the preparation, review or approval of the manuscript; or the decision to submit the manuscript for publication.

