## Supplementary Figures for "Parkinson’s disease genetic risk converges on Lewy body pathology-associated microglial states"

Supplementary Figure 1. Marker and PD risk gene expression across cell types and myeloid states
Supplementary Figure 2. Myeloid state abundance across datasets and pathology, and cross-dataset replication of state structure
Supplementary Figure 3. Pathways enriched in myeloid whole-cell versus nuclear RNA in matched donors
Supplementary Figure 4. GO:BP categories for upregulated marker genes of each myeloid state
Supplementary Figure 5. GO:BP categories for downregulated marker genes of each myeloid state
Supplementary Figure 6. Technical and biological covariates of myeloid state abundances by dataset
Supplementary Figure 7. Cell-cycle pathway enrichment of myeloid state markers
Supplementary Figure 8. Myeloid state abundance changes across Braak LB stages
Supplementary Figure 9. Concordance between IHC and AION-myeloid sn/scRNA-seq proportions of proteostatic-stress microglia across SN neuron loss grades
Supplementary Figure 10. Regional myeloid state abundances and region-specific changes
Supplementary Figure 11. Sex- and age-dependent myeloid state abundance changes
Supplementary Figure 12. Glycolytic, heat shock, and integrated stress response module scores across Braak LB stages
Supplementary Figure 13. ORA of state-specific pathology-associated differentially expressed genes
Supplementary Figure 14. Pathway-level concordance between iPD monocytes and AION-myeloid
Supplementary Figure 15. Spectra factor scores (donor-level collapsed gene programs) across Braak LB stages resolved by brain myeloid state.
Supplementary Figure 16. Heatmap of Parkinson's disease (PD) risk genes' Spectra factor loading scores.

Supplementary Figure 17. Heatmap of Parkinson’s disease (PD) *cis*-eQTL-allele effects on myeloid state abundance

**Supplementary Tables**

Supplementary Table 1. Global differential gene expression analysis results.
Supplementary Table 2. State-specific differential gene expression analysis results.
Supplementary Table 3. De-identified sample metadata.
Supplementary Table 4. Spectra factor association with pathology testing results.
Supplementary Table 5. Spectra factor association across pathology-by-brain-region interactions.
Supplementary Table 6. PD SNP panel differential abundance results (ref vs alt homozygous groups only).
Supplementary Table 7. PD SNP panel differential abundance results (additive model).

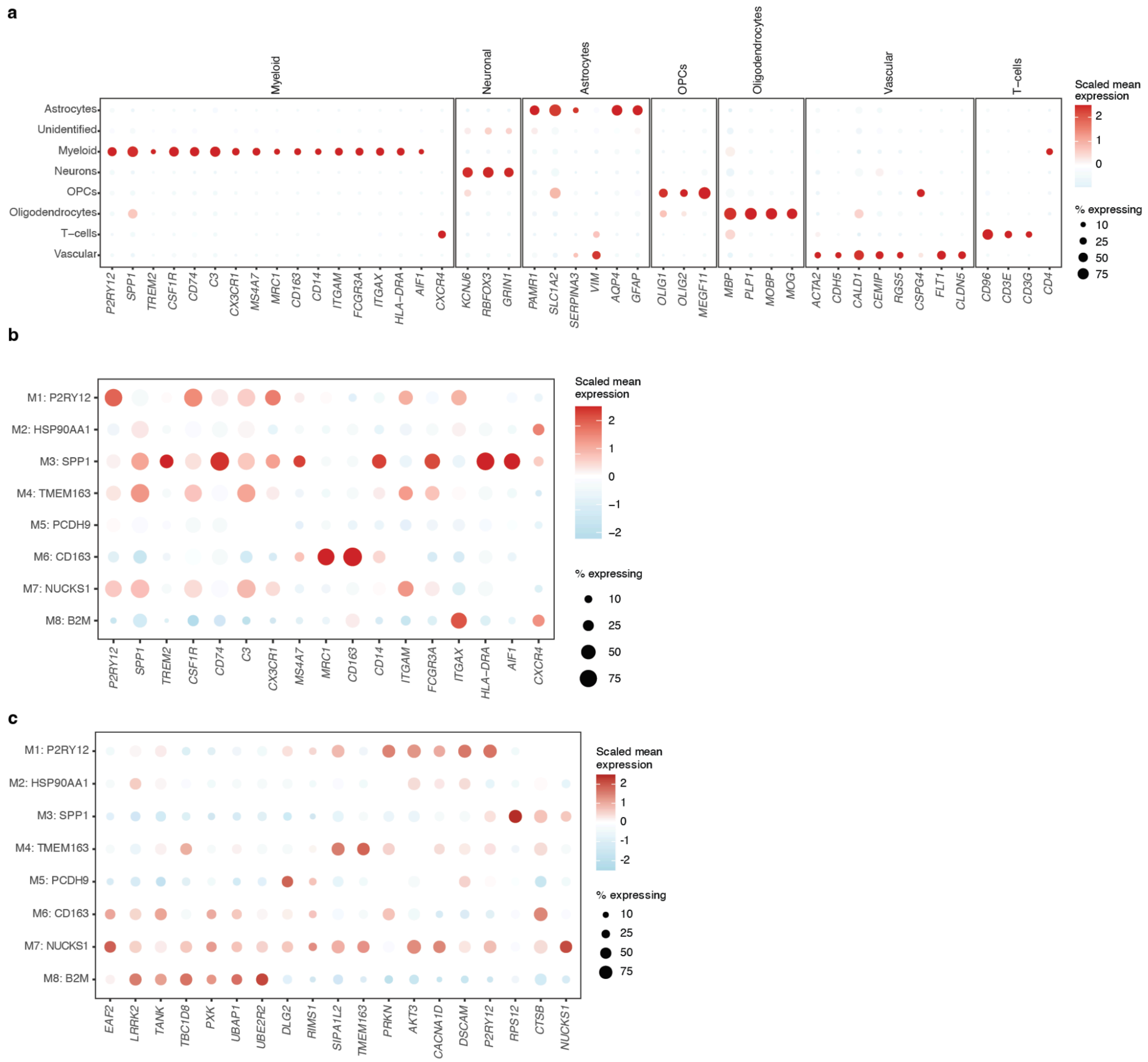


**Supplementary Figure 1. (a)** Dotplot of canonical cell-type marker genes across annotated cell types in all nuclei (n = 423,445). Markers are grouped by lineage: myeloid, neuronal, astrocytes, OPCs, oligodendrocytes, vascular (SMCs, pericytes, endothelial) and T cells. **(b)** Dotplot of pan-myeloid and microglial marker genes across myeloid states (M1-M8) in the myeloid subset (n = 298,592). **(c)** Dotplot of PD genetic risk genes overlapping myeloid state signatures across myeloid states (M1-M8).

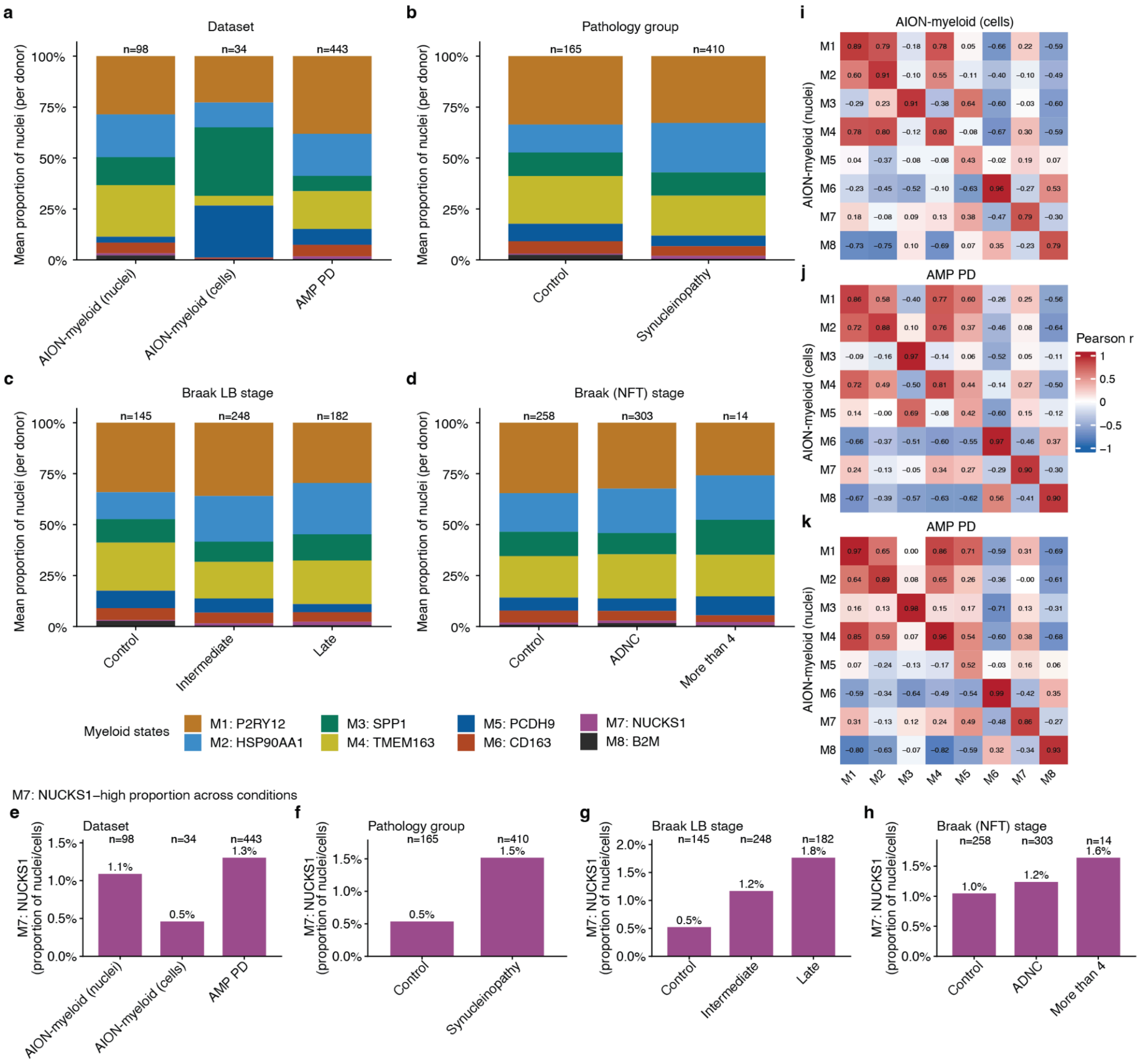


**Supplementary Figure 2. (a-d)** Fractional abundance of each state across datasets and pathology metrics. **(e-h)** M7-NUCKS1-high proportions across dataset, pathology group, Braak LB stage, and Braak NFT stage covariates. **(i-k)** Replication of state structure across AION-myeloid (nuclei), AION-myeloid (cells), and AMP PD myeloid nuclei. Top 50 state markers expression (vst-normalized) was used to calculate cross-dataset correlation structure (Pearson r).

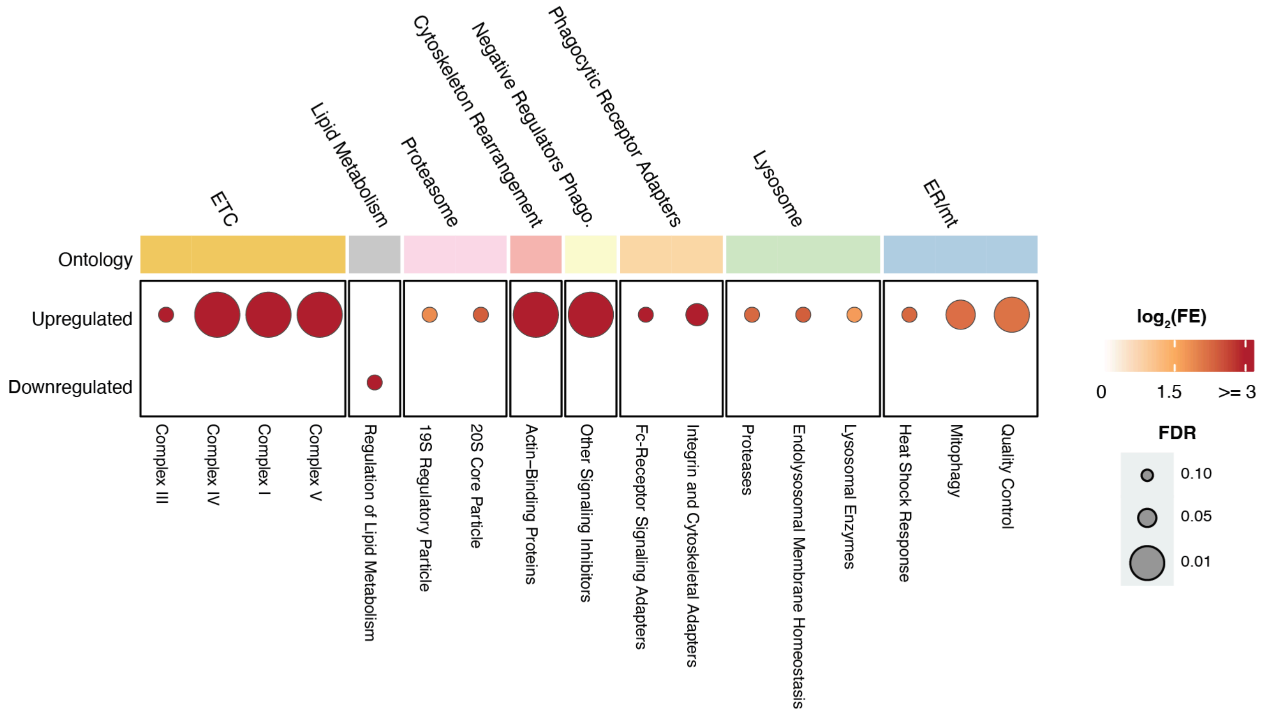


**Supplementary Figure 3.** Heatmap of ORA results testing for myeloid cellular vs nuclear RNA content in a matched-donor analysis (pseudobulk across all states). Pathways (taken from Mattei et al.[1]) shown are upregulated in myeloid cells over nuclei. Dot sizes indicate FDR of enrichment.

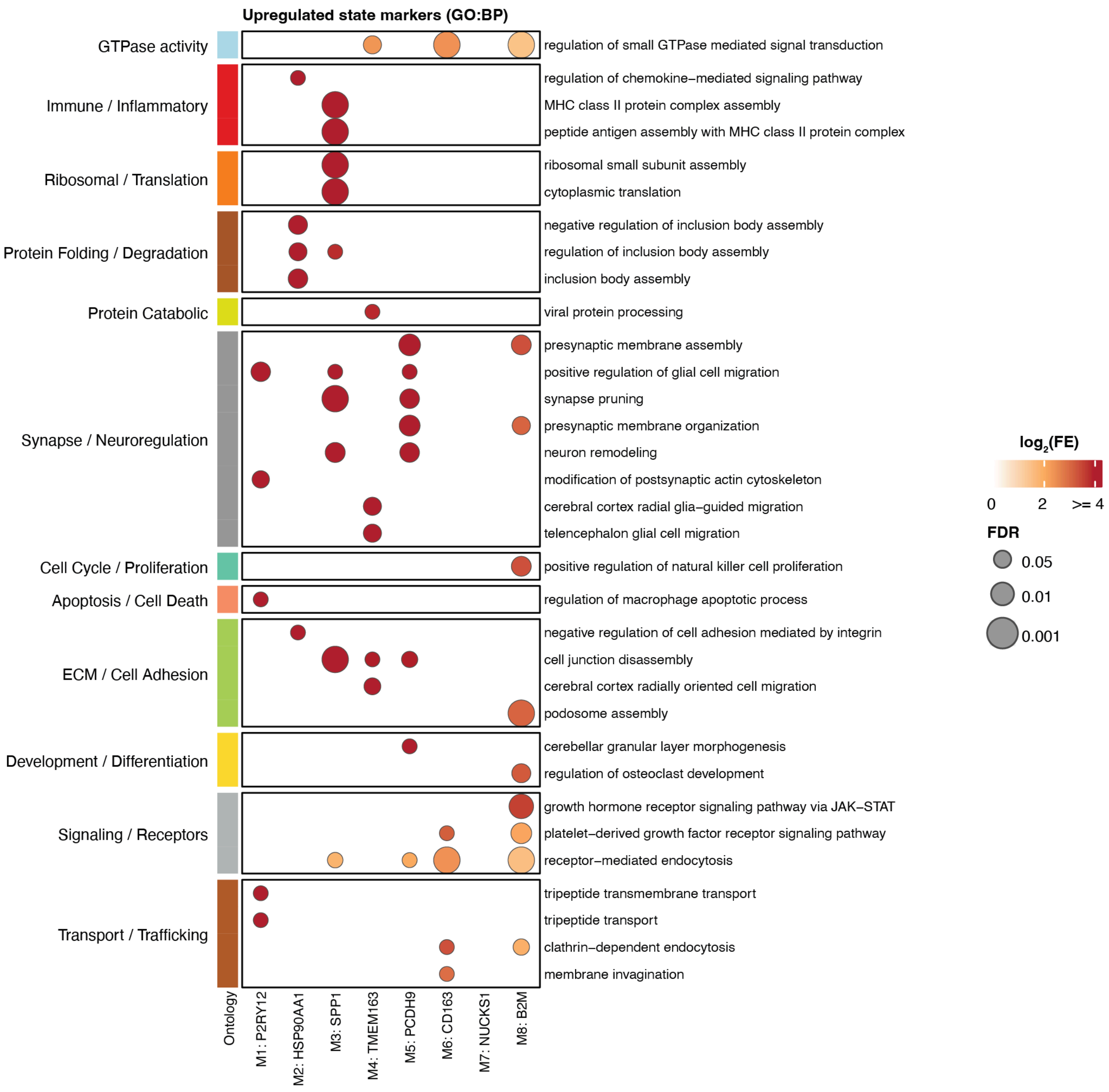


**Supplementary Figure 4.** Top 5 GO:BP functional categories associated with upregulated marker genes of each myeloid state. Each pathway is binned into a supercategory (ontology). Dot sizes indicate FDR of enrichment.

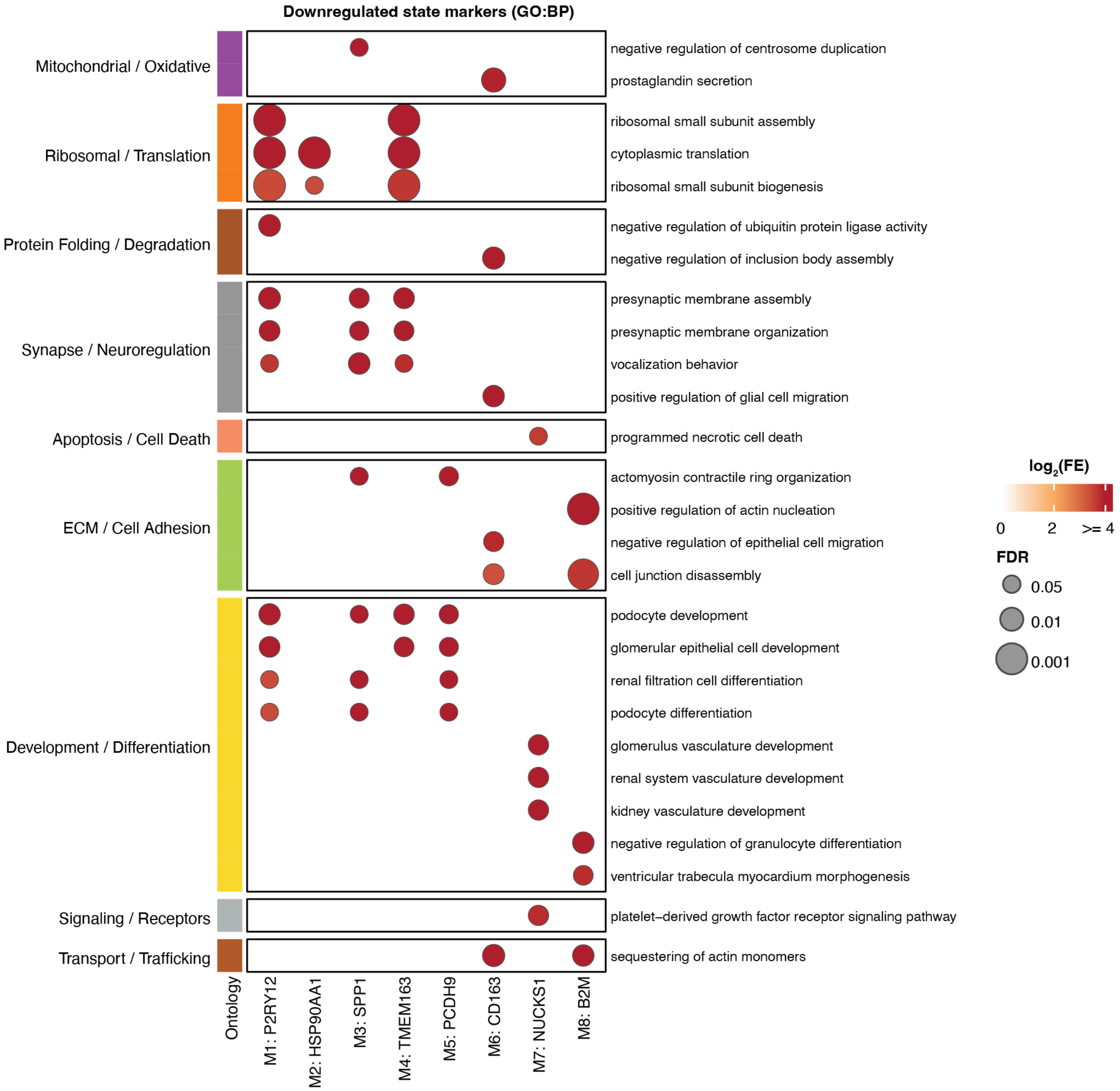


**Supplementary Figure 5.** GO:BP functional categories associated with downregulated marker genes of each myeloid state. Each pathway is binned into a supercategory (ontology). Dot sizes indicate FDR of enrichment.

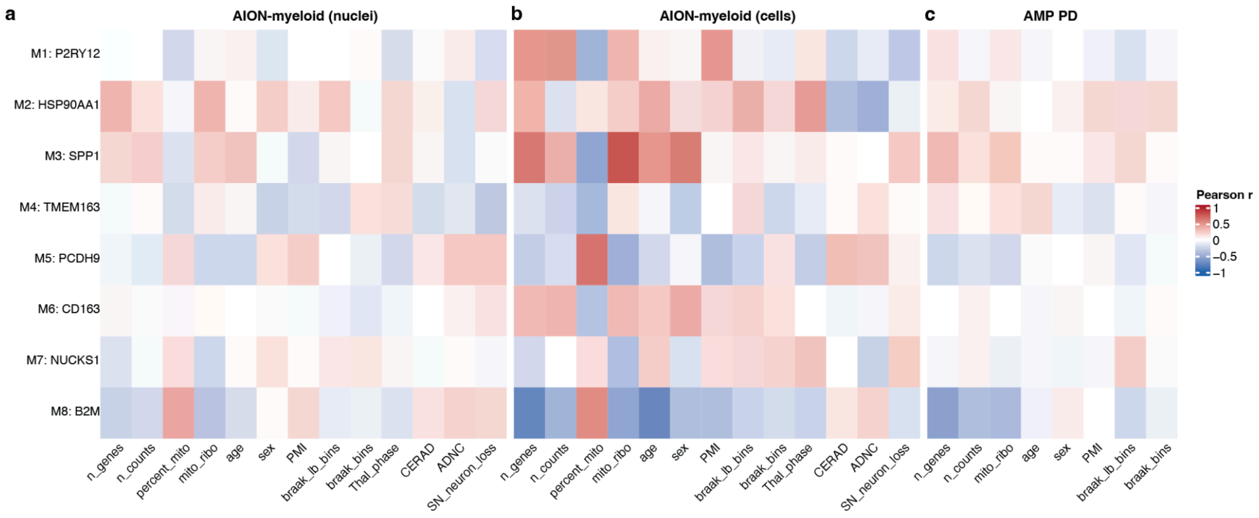


**Supplementary Figure 6. (a-c)** Heatmap of technical and biological covariates for the myeloid state abundances by dataset. Color indicates strength and direction of Pearson's r: red = higher correlation between covariate and state abundance. Blue = lower correlation between covariate and state abundance.

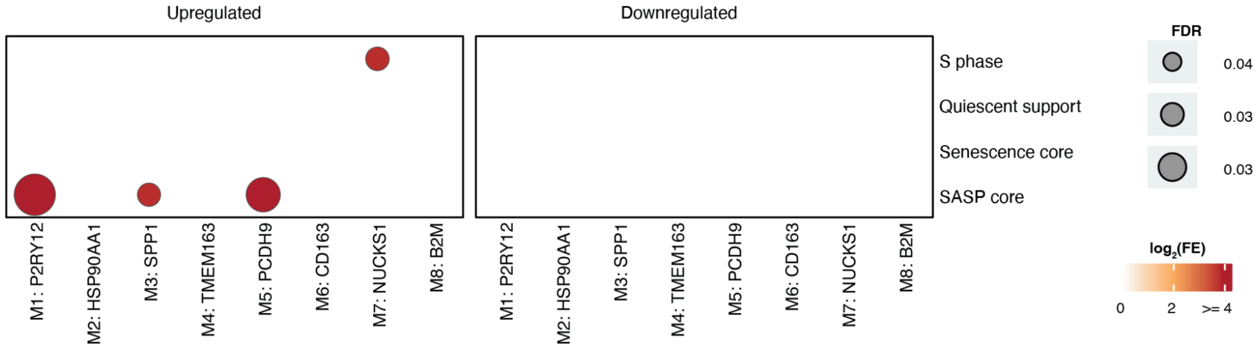


**Supplementary Figure 7.** ORA results of myeloid state markers on cell-cycle-associated gene pathways. Dot sizes indicate FDR of enrichment.

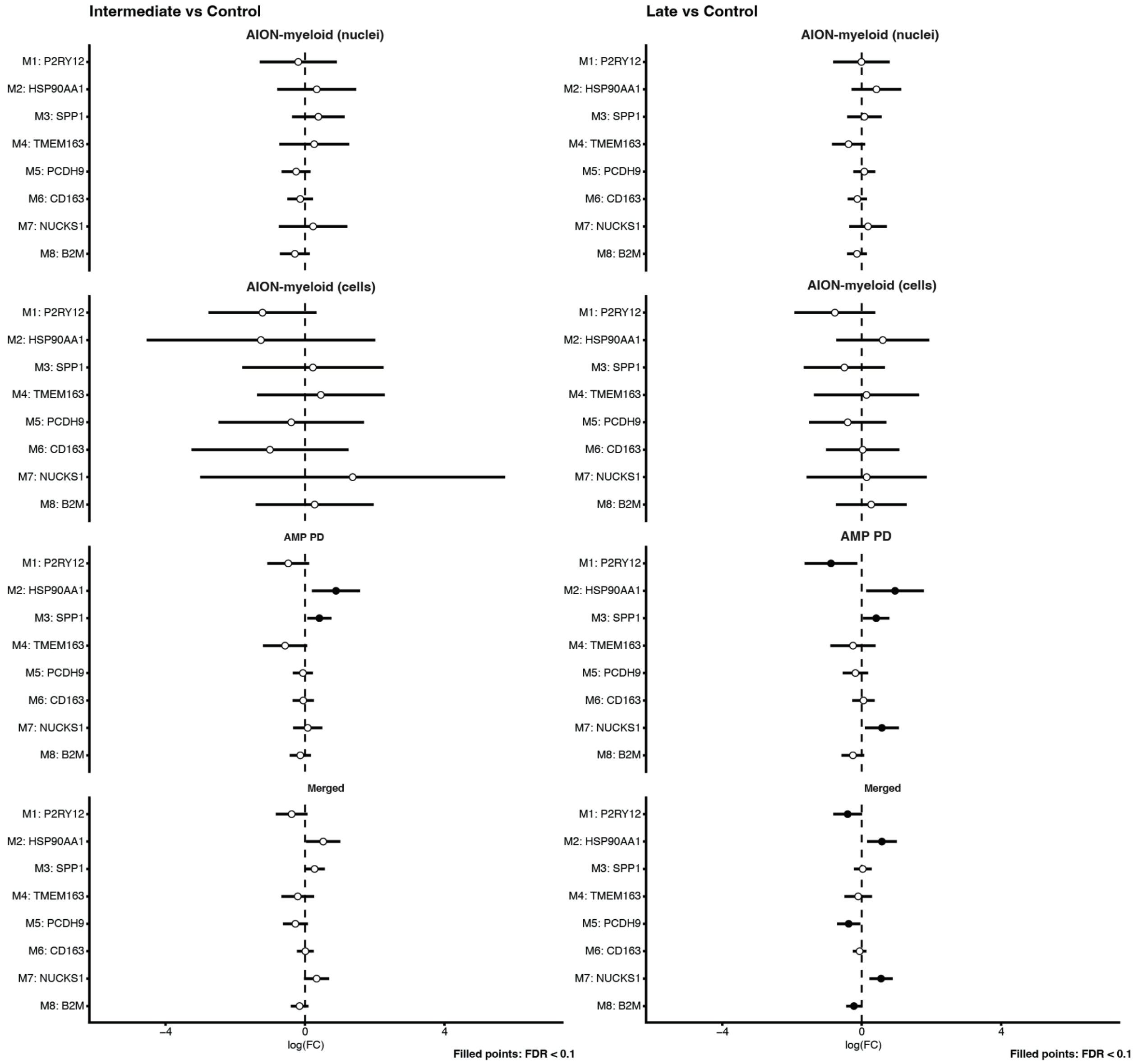


**Supplementary Figure 8.** Crumblr forest plots of abundance differences across Braak LB stages. Filled points denominate significant abundance changes at FDR < 0.1.

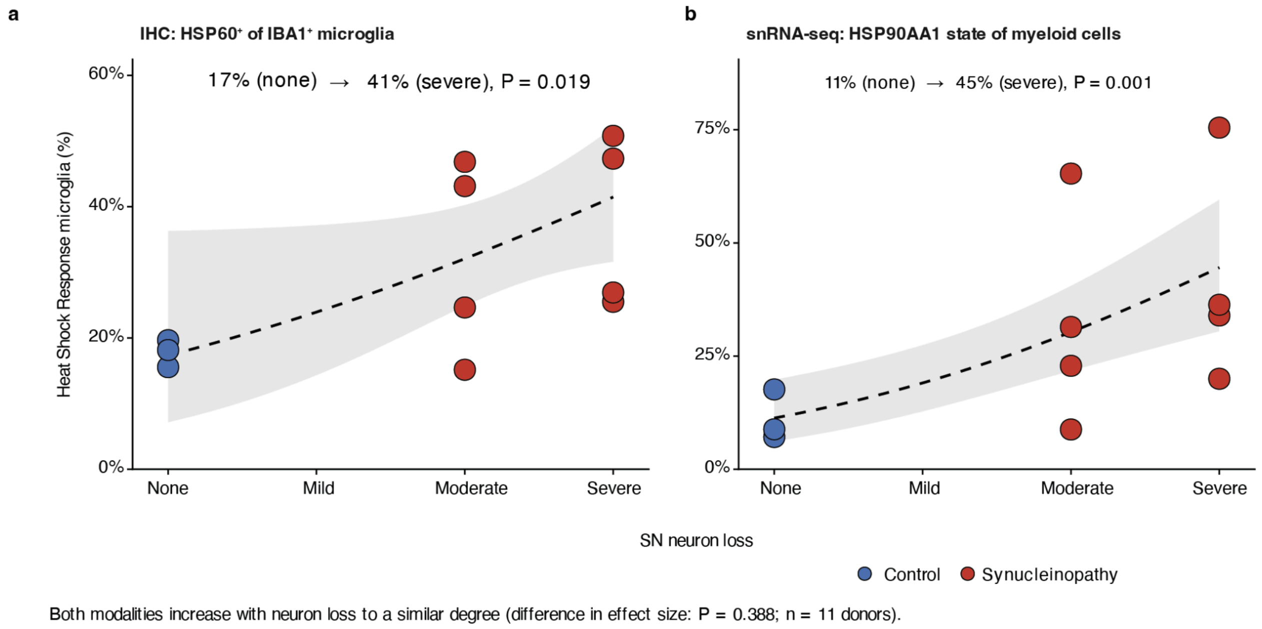


**Supplementary Figure 9. (a)** Proportion of IBA1⁺ microglia co-expressing HSP60 in SN, measured by IHC, by SN neuron-loss grade. **(b)** Proportion of myeloid cells in the M2: HSP90AA1-high microglia state in SN, measured by sn/scRNA-seq (AION-myeloid), by SN neuron-loss grade. Each point is one donor (n = 11). Dashed lines show the fitted trend with 95% confidence bands.

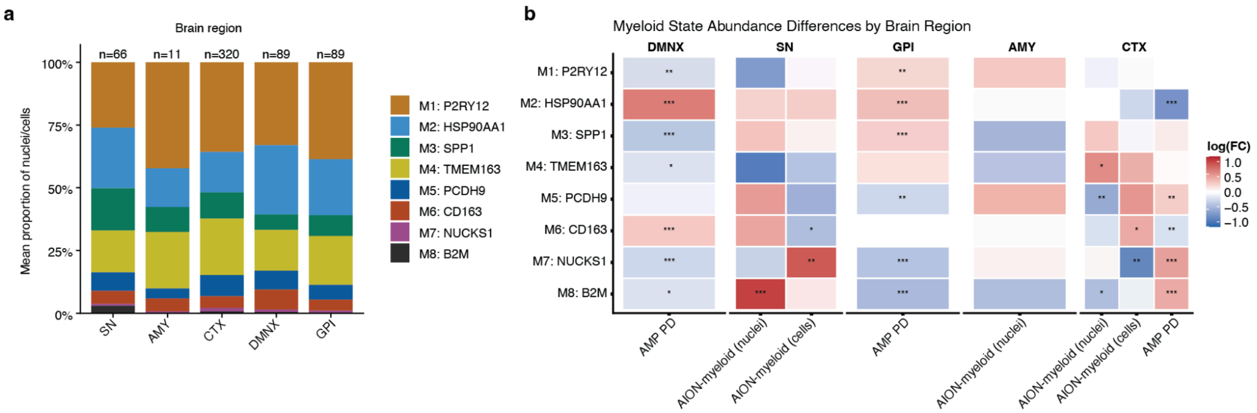


**Supplementary Figure 10. (a)** Regional myeloid state abundances. **(b)** Region-specific abundance changes, highlighting early-affected regions with preferential HSP90AA1-high expansion. ***FDR < 0.01, **FDR < 0.05, *FDR < 0.1

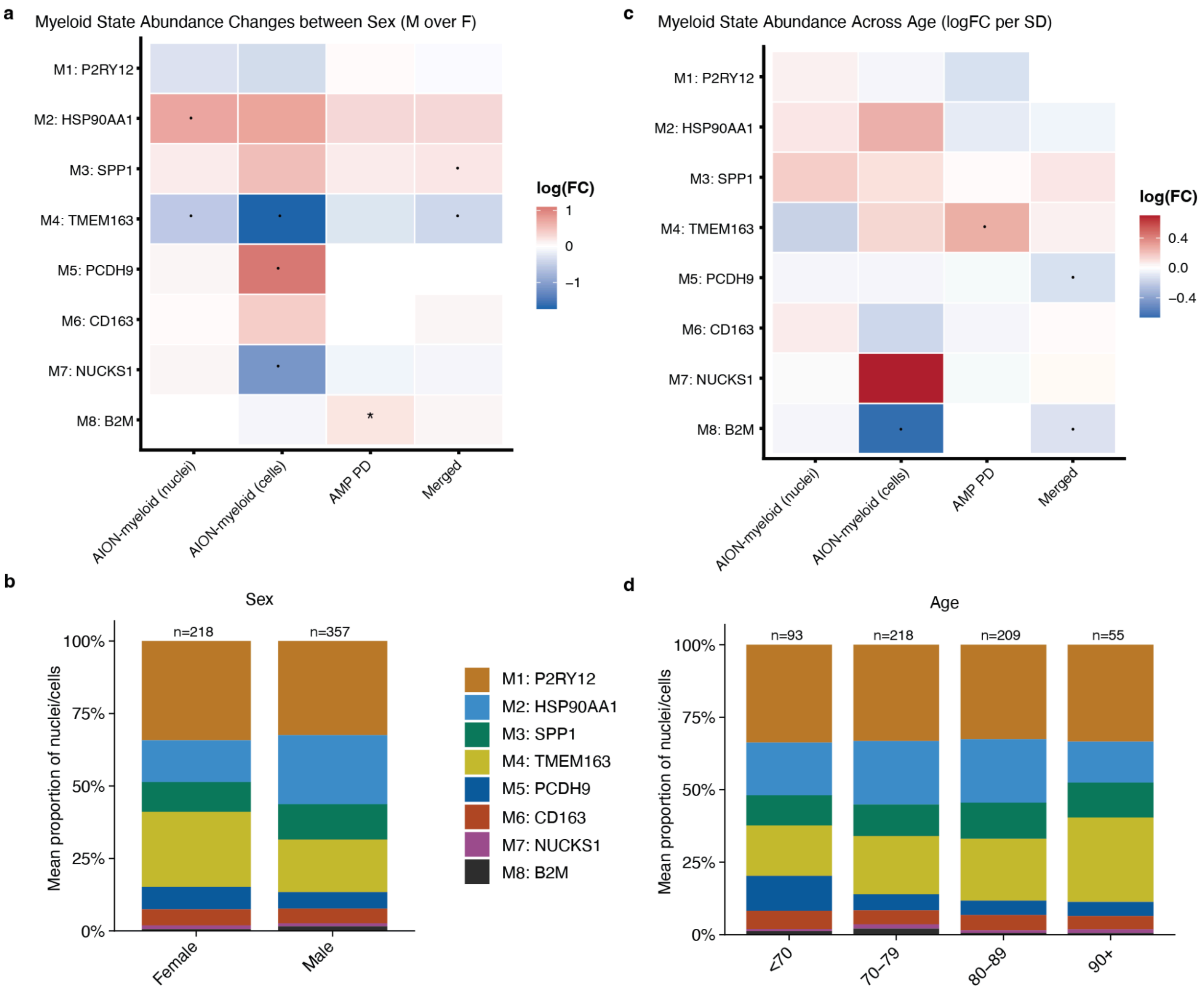


**Supplementary Figure 11. (a-b)** Sex- and **(c-d)** age-dependent abundance changes. Dots indicate $P < 0.05$ of abundance change.

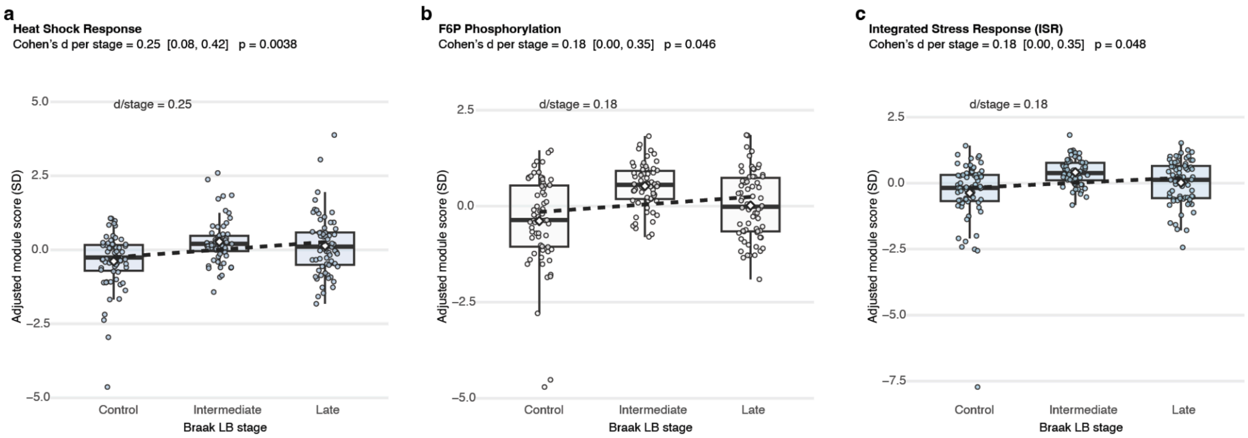


**Supplementary Figure 12.** Donor-level module score (i.e., z-scored mean of that pathway's genes) of glycolytic (**a**) Heat Shock Response (HSR; **b**) and Integrated Stress Response (ISR; **c**) programs across Braak Lewy Body (LB) stages.

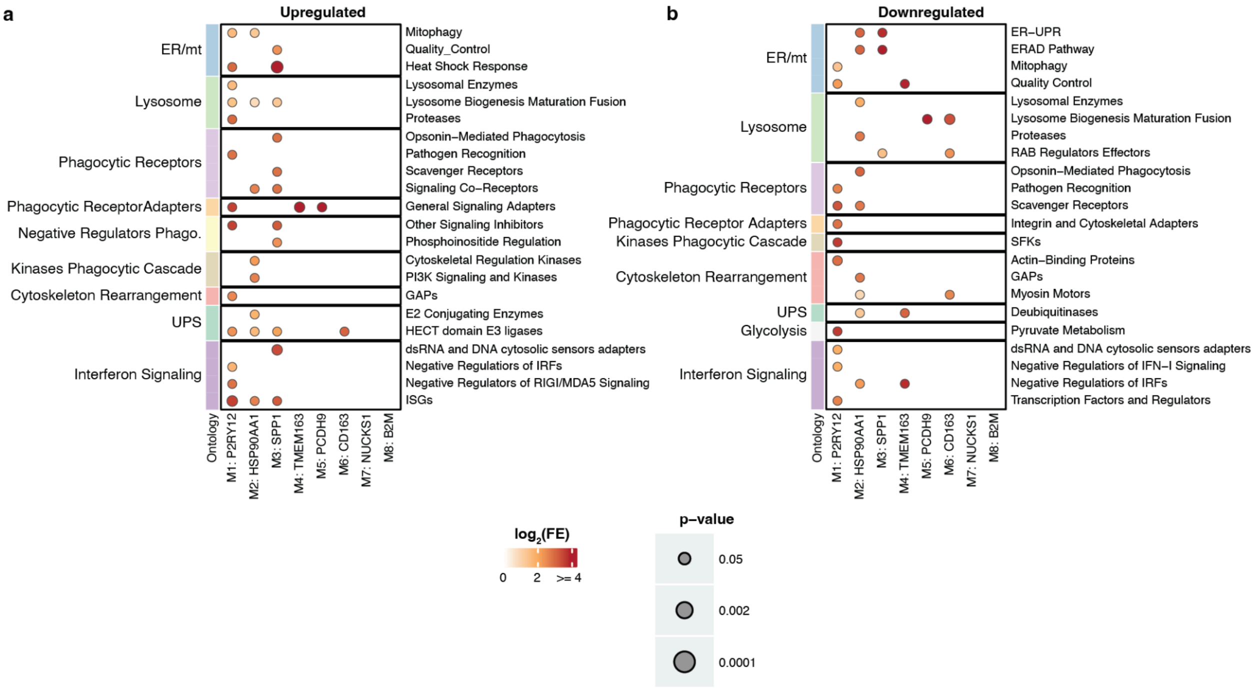


**Supplementary Figure 13.** ORA results of state-specific upregulated **(a)** and downregulated **(b)** differentially expressed genes from a state-dependent pathology vs control differential expression analysis.

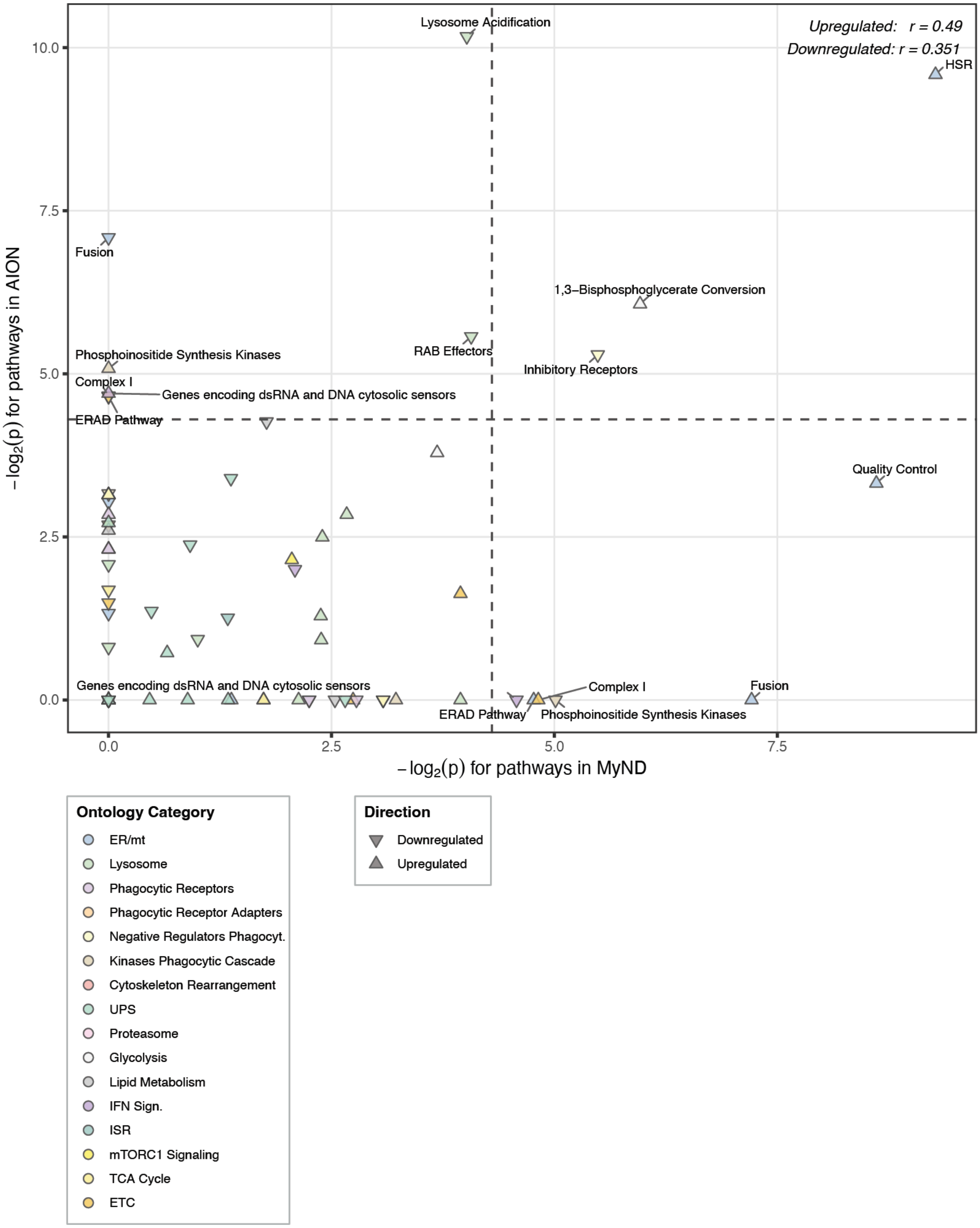


**Supplementary Figure 14.** ORA pathway-level -$\log_2$(p-value) correlation between findings from Control vs iPD monocytes from Mattei et al.[1] (x-axis) and AION-myeloid (y-axis). Myeloid cells in Neurodegenerative Diseases (MyND).

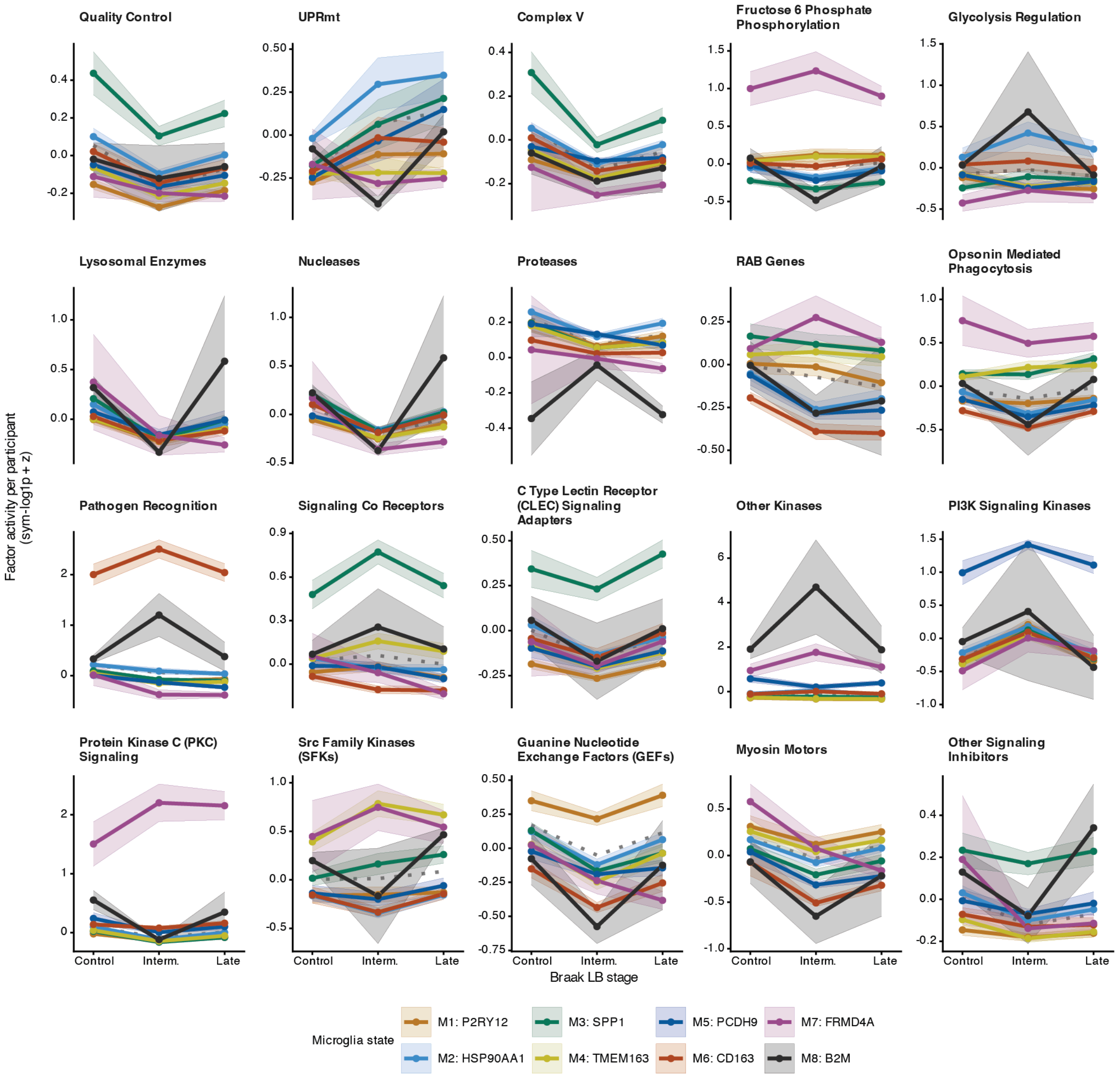


**Supplementary Figure 15.** Spectra factor scores (donor-level collapsed gene programs) across Braak LB stages resolved by brain myeloid state. The dotted gray line shows the mean across all states. Activity is the z-scored, cell-level factor score (s.d. units). Braak LB stages: Control, 0; Intermediate, 1-4; Late, 5-6. Statistics are given in the main text and **Supplementary Tables 4-5**.

**PD risk gene Spectra loadings per factor supercategory**

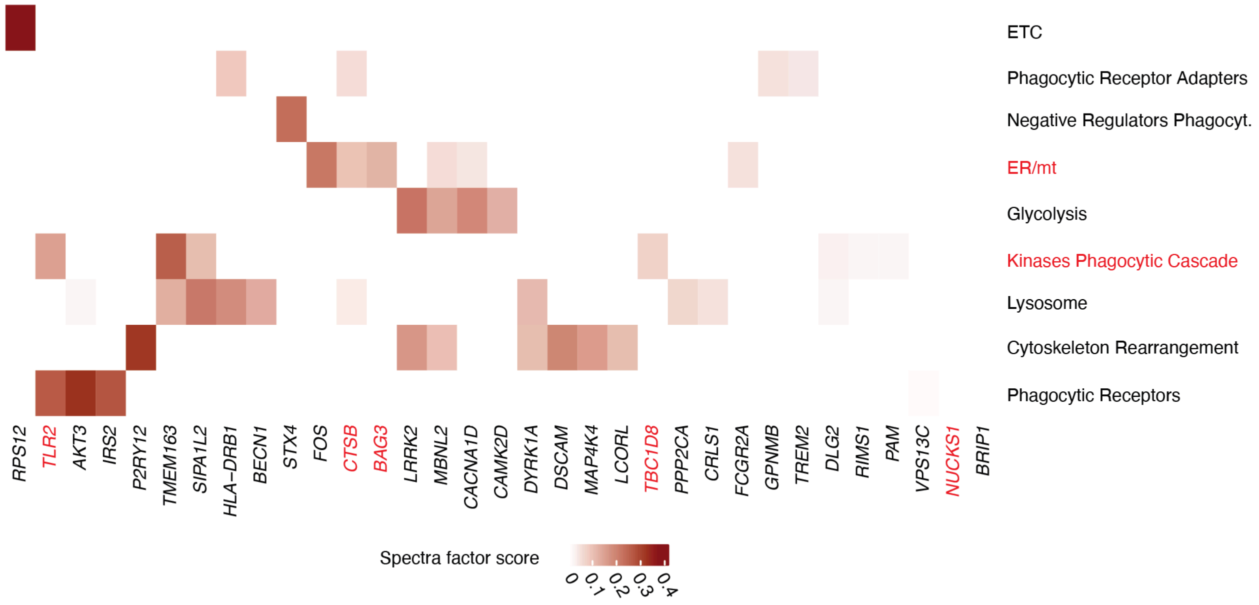


**Supplementary Figure 16.** Heatmap of Parkinson's disease (PD) risk genes' Spectra factor loading scores. The factors' super-ontology is on the y-axis. On the x-axis are PD risk genes. Supercategories in red are enriched for PD polygenic risk in the scDRS results. Genes in red are discussed in the text as state-enriched PD risk genes

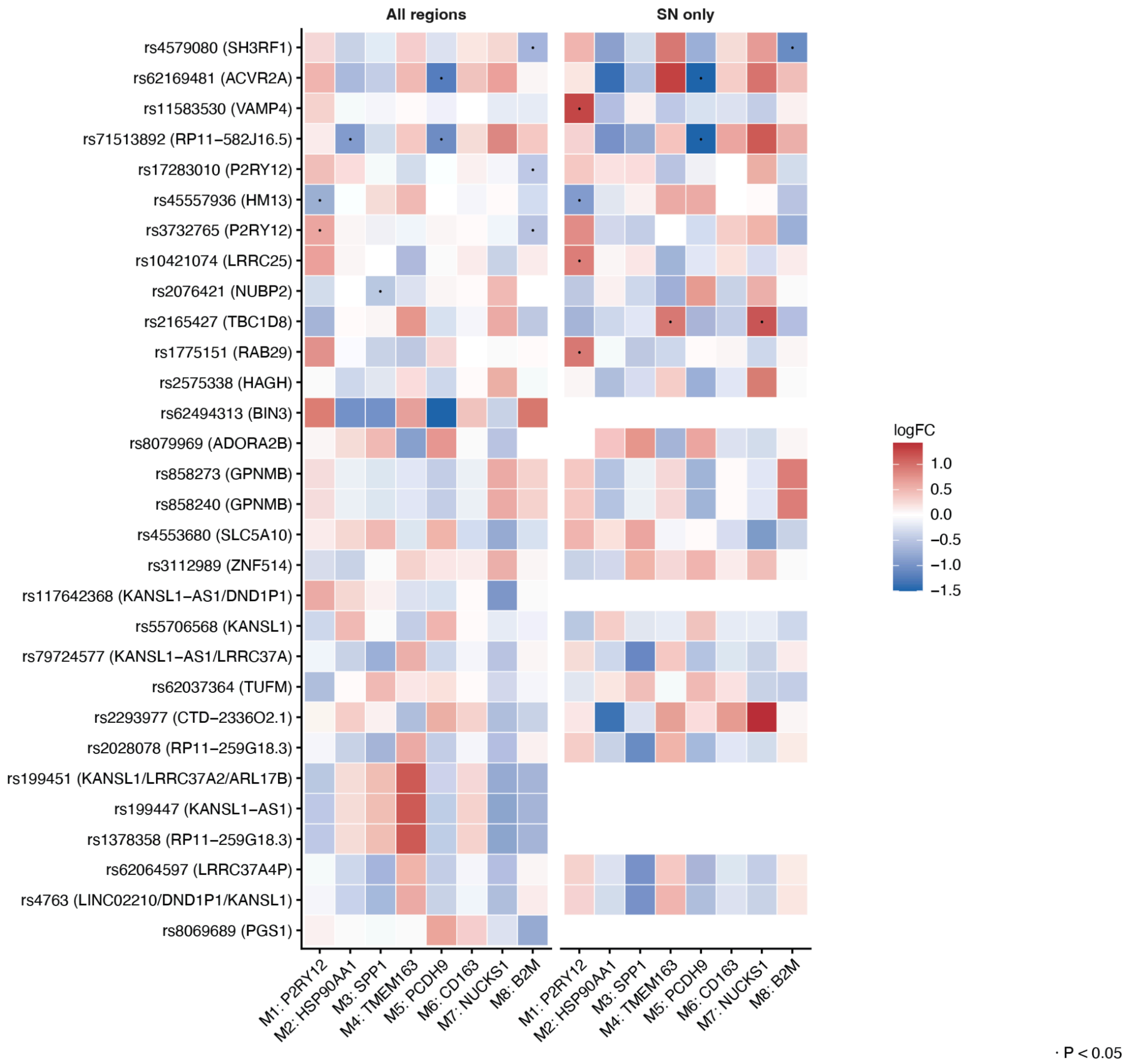


**Supplementary Figure 17.** Heatmap of Parkinson's disease (PD) *cis*-eQTL-allele effects on myeloid state abundance. Rows show PD-associated microglial *cis*-eQTL variants and columns show myeloid states split by brain-region-dependent analysis (all regions and SN-only). Tile color indicates the difference in centered log-ratio (CLR)-transformed state abundance between homozygous alternate and homozygous reference donors. Red indicates higher relative abundance per additional effect allele and blue indicates lower. Dots denote $P < 0.05$.
